# Integrated surveillance resolves Darién paradox of Oropouche virus emergence in Panama’s migration corridor

**DOI:** 10.64898/2026.05.28.26354376

**Authors:** Xacdiel Rodríguez, Juan G. Perez-Jimenez, Laura W. Alexander, Carlos Lezcano-Coba, Josefrancisco Galué, Yelissa Juárez, Davis Beltrán, Darci R. Smith, Malik Kadir, Danielle W. Ali, Rita Corrales, Lidimarie Trujillo Rodriguez, Ghyssella E. Valdiviezo, Quinn K. Thomas, Anthony Cicalo, Maren Fitzpatrick, Andrea Luquette, LT. Cameron Sayer, Regina Z. Cer, Francisco Malagon, Ilka Anabel Grajales, Luis Felipe Rivera, Zuleyka González-R, Juan Antioco, Ellianys Walters-Valdes, Niccolò Meneghello-Ponce, Amy Y. Vittor, Kiriam Escobar-Lee, Aaron Abouganem-Shaw, Fátima Rodríguez, Eduardo Aguirre, Steev Loyola, Yeny Tinoco, Brechla Moreno, María Chen-Germán, Sonia Ampuero, Adrean Gómez-Angelo, Samir Correa-Duarte, José Acevedo, Blas Ramos, Maria Eugenia de Antinori, Claudia Gonzalez, Oris Chavarria, Jessica Gondola, Ambar Moreno, Celestino Aguilar, Pablo Gonzáles, Carmela Jackman, Hector Cedeño, Bernardo Gutiérrez, Moritz U.G. Kraemer, Victor Saldaña, Rodrigo DeAntonio, Alexander A Martinez, Blas Armién, Juan Miguel Pascale, Arlene Calvo, Mauricio L. Nogueira, William M. de Souza, Kathryn A. Hanley, Nuno R. Faria, Ilaria Dorigatti, Nikos Vasilakis, Christl A. Donnelly, Sandra López-Vergès, Kimberly A. Bishop-Lilly, Carla Mavian, Ana I. Bento, Jean-Paul Carrera

## Abstract

Oropouche virus (OROV) spread across the Americas in 2024, yet Panama’s Darién migration corridor saw no outbreak until nearly a year after Brazil’s January 2024 peak, raising two hypotheses: cryptic circulation masked by diagnostic gaps, or recent introduction under permissive climatic conditions. Here we resolve this paradox using integrated clinical, genomic, and climate-informed surveillance. Among 1,040 individuals tested, 43% were OROV-positive and showed a clinical signature distinct from co-circulating arboviruses, including headache more frequent than in dengue (RR 2.38, 95% CI 1.74-3.24). The household secondary attack rate was 56%, and waste burning independently predicted infection. Phylogeographic reconstruction identified a single recent introduction in October 2024 with no evidence of adaptive evolution, excluding prolonged cryptic persistence. Climate-informed models indicate broad outbreak susceptibility across Panama, with Bocas del Toro and Los Santos as the next highest-risk provinces. These findings identify a Central American foothold for OROV with potential for further northward spread.

## Main

In 2024, Oropouche virus (OROV) surged across the Americas, with >10,000 laboratory-confirmed cases across more than ten countries and travel-associated infections reported in Europe and North America^1^. As of March 2026, no licensed vaccines, antivirals, or routine surveillance systems exist for OROV^2,3^. OROV is an Orthobunyavirus of the family Peribunyaviridae^4–6^ transmitted by the biting midge *Culicoides paraensis* in sylvatic cycles involving sloths and other mammals^7–9^. It causes an acute febrile illness clinically resembling dengue, Zika, and chikungunya: headache, myalgia, rash, with potential neurological complications; documented vertical and sexual transmission has broadened its threat profile^2,3,5^.

Until 2024, OROV had remained largely confined to South America, with the 1989 Bejuco outbreak^10^ the only previously documented autochthonous transmission in Central America. In Panama, prior serologic evidence and sporadic febrile cases (most recent in August 2024) had been interpreted as consistent with cryptic endemic circulation^11^. The 2024 surge changed this pattern: Brazil alone reported >12,000 cases including the first extra-Amazon autochthonous transmission, driven by a reassortant strain (OROV_BR-2015-2024_) implicated in the regional expansion^12–14^. Broad routine surveillance for OROV remains absent across the region^13^.

Recent genomic^12,14^ and ecological modeling^15,16^ studies have defined OROV’s evolutionary dynamics and transmission drivers in Brazil. Key gaps persist in Central America. The timing and source of introductions remain unresolved. It is unclear whether Panama represents an endemic foothold or a recent incursion. Whether OROV has clinical or immunologic signatures that can improve case-finding in dengue-endemic settings is unknown. And whether Brazil-calibrated risk models prospectively identify high-risk regions in new ecological contexts has not been tested. The delayed emergence (November-December 2024) in Darien, Panama’s high-risk migration corridor (∼500,000 annual human crossings^17^), where no outbreak was reported until nearly one year after Brazil’s January 2024 surge, presents a paradox with two competing explanations: either OROV was already circulating cryptically, undetected because undifferentiated febrile illness is routinely attributed to dengue; or OROV was introduced de novo during a window of climatic permissiveness in late 2024. This distinction carries direct implications for surveillance design and for whether Panama represents a new foothold for northward spread into Mesoamerica. Panama’s environment combines *C. paraensis* distribution with extreme mobility, creating ideal arbovirus seeding conditions; local health systems default to dengue diagnosis for undifferentiated fever, masking OROV circulation through inadequate molecular surveillance^18^. Viral seeding timing, aligned with climate and mobility windows, likely governs the likelihood of sustained transmission^11,15^.

In this study, we investigated the 2024–2025 Darién outbreak using integrated surveillance (1,040 tested; 447 confirmed cases; 43% positivity), clinical profiling (compared to co-circulating arboviruses), genomic reconstruction (new 34 genomes for L and M segments, and 29 for S segment), and risk models trained on Brazilian surveillance data using outbreaks beginning from July 2021 to December 2024 to resolve cryptic transmission vs. recent introduction. We show that the delayed outbreak reflects a single introduction in October 2024 from a Cuba-linked ancestor enabled by anomalous climatic conditions, excluding prolonged cryptic persistence; we identify a syndromic and cytokine signature that distinguishes OROV from dengue and chikungunya to guide diagnostics in dengue-endemic settings; document a 56% household secondary attack rate alongside substantial asymptomatic burden; and demonstrate that climate-informed models predict outbreak-permissive conditions across much of Panama (mean modeled susceptibility 56%), with Bocas del Toro and Los Santos as next highest-risk provinces. These findings reveal OROV’s Central American foothold, expose diagnostic blind spots in dengue-endemic surveillance, and define urgent containment priorities ahead of northward migration threats.

## Results

### The initial OROV outbreak in Darién, Panama

Following reports of an unusual increase in febrile illness in Darién Province, Panama, between 24 and 25 December 2024, an epidemiological investigation was initiated. Between 20 December 2024 and 14 March 2025, a total of 1,040 individuals (including 133 asymptomatic contacts) were evaluated for OROV infection in Darién Province, Panama (Figure 1; more detail in Supplementary Materials Figure S19 and Table S15). Surveillance was conducted through three complementary strategies: routine surveillance at health care facilities (n = 690), community-based field investigations (n = 292), and enrollment through telephone outreach (n = 58). Overall, 447/1040 laboratory-confirmed OROV infections/recent exposures were identified, corresponding to an overall positivity of 43.0% (95% CI, 40.0–46.0).

**Figure 1.**
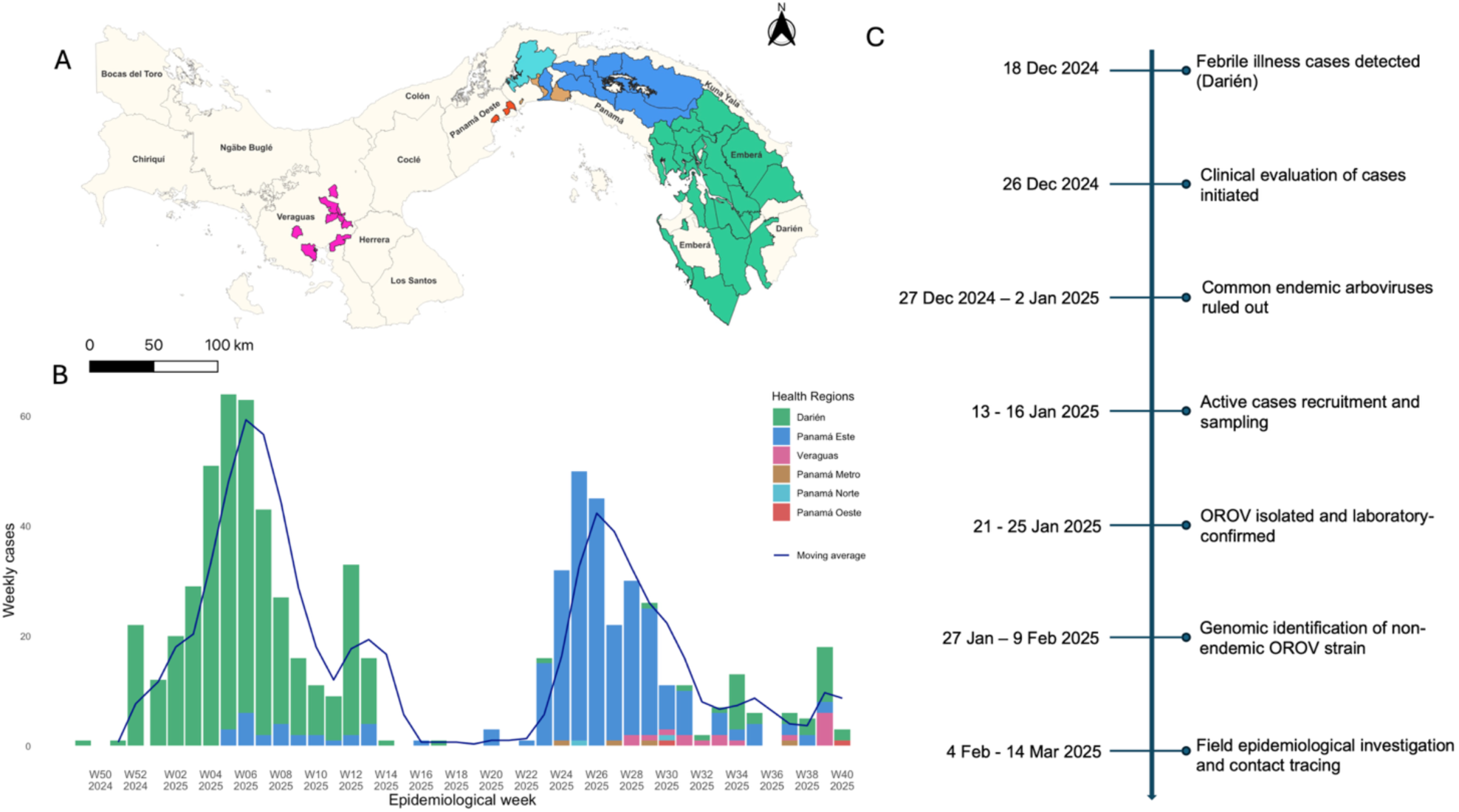
Oropouche virus and spread in Panama (December 18, 2024–October 26, 2025). **A)** Map of Panama with OROV reported cases. Regions that reported cases are in color. **B)** Cases reported in Panama for the period of December 18, 2024–October 26, 2025. Colors match the regions in panel A. Moving average depicted in black line. **C)** Darién outbreak investigation timeline.

During routine clinical surveillance between 20 December 2024 and 14 March 2025, 690 individuals presenting with undifferentiated febrile illness were evaluated at primary healthcare facilities, of whom 214 were laboratory-confirmed as OROV infections, yielding a positivity rate of 30.9% (95% CI, 27.5–34.5). Most infections in this group were identified by RT-PCR (148/214; 69.2%; 95% CI, 62.5–75.3) at median of one day after symptom onset (IQR: 0–2), followed by anti-OROV IgM ELISA (64/214; 29.9%; 95% CI, 23.9–36.5) at a median of 10.5 days after symptom onset (IQR: 7–16), with a small proportion confirmed by both methods (2/214; 0.9%). A field epidemiological investigation was conducted in communities reporting the highest number of cases. During household visits, residents were invited to participate voluntarily; individuals younger than 4 years or older than 70 years were excluded according to the approved bioethics protocol. Between 14 January and 27 February 2025, we enrolled 292 participants, including both individuals with current or recent symptoms compatible with arboviral infection and asymptomatic individuals. Among those reporting at least one symptom, 104 of 156 participants tested positive for OROV (66.7%; 95% CI, 58.7–74.0), with infections predominantly identified by IgM ELISA (79/104; 76.0%) at median of seven days after symptom onset (IQR: 2-–22), followed by combined IgM/IgG ELISA (11/104; 10.6%), RT-PCR (9/104; 8.6%) at a median of one day after symptom onset (IQR: 0–2), IgG/PRNT (3/104; 2.9%) at a median of 12.5 days after symptom onset (IQR 5–31.5), and RT-PCR/IgM (2/104; 1.9%). Notably, 73 of 136 asymptomatic participants also tested positive (53.8%). Among these participants, evidence of recent OROV infection was primarily detected by IgM ELISA (70/73; 95.9%), whereas active infection was confirmed by RT-PCR in a smaller proportion of cases (3/73; 4.1%), supporting the occurrence of inapparent OROV infections within the community.

In addition, a contact-based telephone enrollment conducted between 18 and 25 February 2025 reached out to individuals with a documented history of febrile illness with symptom onset between December 2024 and February 2025. Among the 58 participants who consented, 56 had laboratory-confirmed OROV infections (96.6%; 95% CI, 88.1–99.6), with most cases confirmed by RT-PCR (55/56; 98.2%) at a median of one day (IQR: 0–2) and ELISA IgM (1/56; 1.2%) at a median of 74 days and symptom onset dates extending back to 8 December 2024.

Across all surveillance components, OROV infections were detected more frequently among women (255/447; 57.0%), and adults 18–59 years old represented the most affected age group (305/447; 68.2%), followed by older adults ≥60 years old (53/447; 11.9%). Case confirmation relied primarily on RT-PCR (215/447; 48.1%; median one day, IQR: 0–2) and IgM ELISA (214/447; 47.9%; median 10 days, IQR: 5–19), whereas fewer cases were identified by combined IgG ELISA and neutralization assays at a later stage (3/447; 0.7%; median 12.5 days, IQR: 5–31.5), as well as by combined IgM/IgG ELISA (13/447; 2.9%) and RT-PCR/IgM (2/447; 0.45%) (Supplementary Materials, Table S14). A total of 75 viral isolates were successfully obtained including 54 from samples that were RT-PCR positive for OROV prior to isolations and 21 that had not previously been tested by RT-PCR but were subsequently confirmed as OROV-positive following viral isolation.

### Geographic distribution of OROV cases

Among permanent residents of Darién, 408 OROV infections were identified between December 2024 and March 2025, through a combination of RT-PCR, ELISA (IgM/IgG), PRNT, and viral isolation. Based on the 2023 provincial census population (54,235 inhabitants), this corresponds to an incidence of 752.3 infections per 100,000 inhabitants (95% CI, 679.1–825.4). Incidence was highest in the districts of Pinogana (1,161.7 infections per 100,000), Santa Fe (400.4 per 100,000) and Chepigana (315.8 per 100,000). Pinogana was identified as the principal focus of transmission within Darién, driven by a high number of reported cases in Metetí (170 OROV confirmed cases) (Supplementary Materials, Figure S19 and Table S15).

In addition, 29 OROV infections (59.0% positivity) were detected among individuals temporarily stationed in Darién (e.g., National Border Service personnel) who reside outside the province, including individuals residing in the following provinces: Panamá (13), Veraguas (7), Panamá Oeste (5), Chiriquí (2) and Coclé (2). A further 10 cases (33.3%) occurred in participants with unknown or unreported communities of residence.

### Acute OROV infection induces early pro-inflammatory and antiviral immune responses

We analyzed 97 serum samples obtained from permanent residents of Darién Province, Panama, including 56 RT-qPCR-confirmed OROV-positive febrile individuals recruited from healthcare centers between December 2024 and March 2025, all presenting with mild disease and ≤7 days since symptom onset, and 41 OROV-negative healthy individuals from a cross-sectional study conducted in Darién in 2018. Negative controls were asymptomatic and tested negative by all laboratory diagnostic assays for OROV and were matched to OROV-positive individuals by age range and sex (Table S16). Most acute samples were collected after one day (24 h) of symptom onset. Viral load and soluble cytokines were quantified. Most samples had a viral load over the limit of quantification of 1000 copies/mL, with a median around 3×10⁷ copies/mL. Though high viral titres (>10⁷ copies/mL) were confined to samples collected within two days of symptom onset (Fig. 2A), there was no statistical differences for viral load associated to days of symptoms onset (p=0.797), sex (p=0.516) or age range (p=0.119). Acute OROV infection elicited a pronounced pro-inflammatory immune signature and a robust antiviral interferon response (Fig. 2B). Infected individuals exhibited markedly elevated levels of the chemokine IP-10 (200.9 pg/mL in OROV-positive vs 2.5 in healthy negatives, p < 0.0001), and significant upregulation was observed for type I IFN-α2 (3584 vs. 1268; p < 0.0001), type II IFN-γ (3504 vs. 1347; p < 0.0001), and type III IFN-λ1 (3311 vs. 1540; p < 0.0001) along with IL-6 (1718 vs. 1209; p = 0.0012). Concurrently, the anti-inflammatory cytokine IL-10 was elevated (2338 vs. 588; p < 0.0001). In contrast, IL-8 levels were significantly reduced during infection (1395 vs. 1531; p < 0.0001). IL-1β (p = 0.0023) and IL-6 levels were also lower (3.89 vs 13.76, P = 0.0012) during OROV acute infection. No significant induction was observed for TNF-α, IL-12p70, GM-CSF, IFN-β, and IFN-λ2 (IL-28A). Among 56 OROV-positive participants included in cytokine analyses, bivariate regression models adjusted for age and sex (Fig. 2C) showed that early acute infection (≤2 days since symptom onset) was weakly positively associated with IP-10 levels (RR: 1.11, 95% CI: 0.99–1.23; P = 0.053), and to a lesser extent with IL-8, IL-6 and viral load quantification (RR: 1.03, 95% CI: 1.00–1.06; P = 0.054), whereas IL-12p70 showed an opposite trend (RR: 0.891, 95% CI: 0.61 – 1.30; p = 0.055). However, none of these associations reached statistical significance and should therefore be considered preliminary findings warranting replication in larger cohorts. Because viral load may be modulated by the immune response, additional bivariate regression analyses adjusted for age and sex evaluated associations between cytokine levels and viral load. Several cytokines implicated in inflammatory and interferon-mediated responses showed positive associations with viral load, although most did not reach statistical significance, including IP-10 (RR: 1.03, 95% CI: –0.07 – 2.12; p = 0.073). In contrast, GM-CSF (–0.87, 95% CI: –1.50 – –0.25; p = 0.009) and IL-12p70 (–0.49, 95% CI: –0.89 – –0.08; p = 0.024) showed significant inverse associations with viral load. Together, these findings suggest that early OROV infection may involve coordinated inflammatory and interferon-related immune activation, while IL-12p70 could be associated with lower viral loads. However, these results should be interpreted with caution given the limited sample size and require validation in larger cohorts.

**Figure 2.**
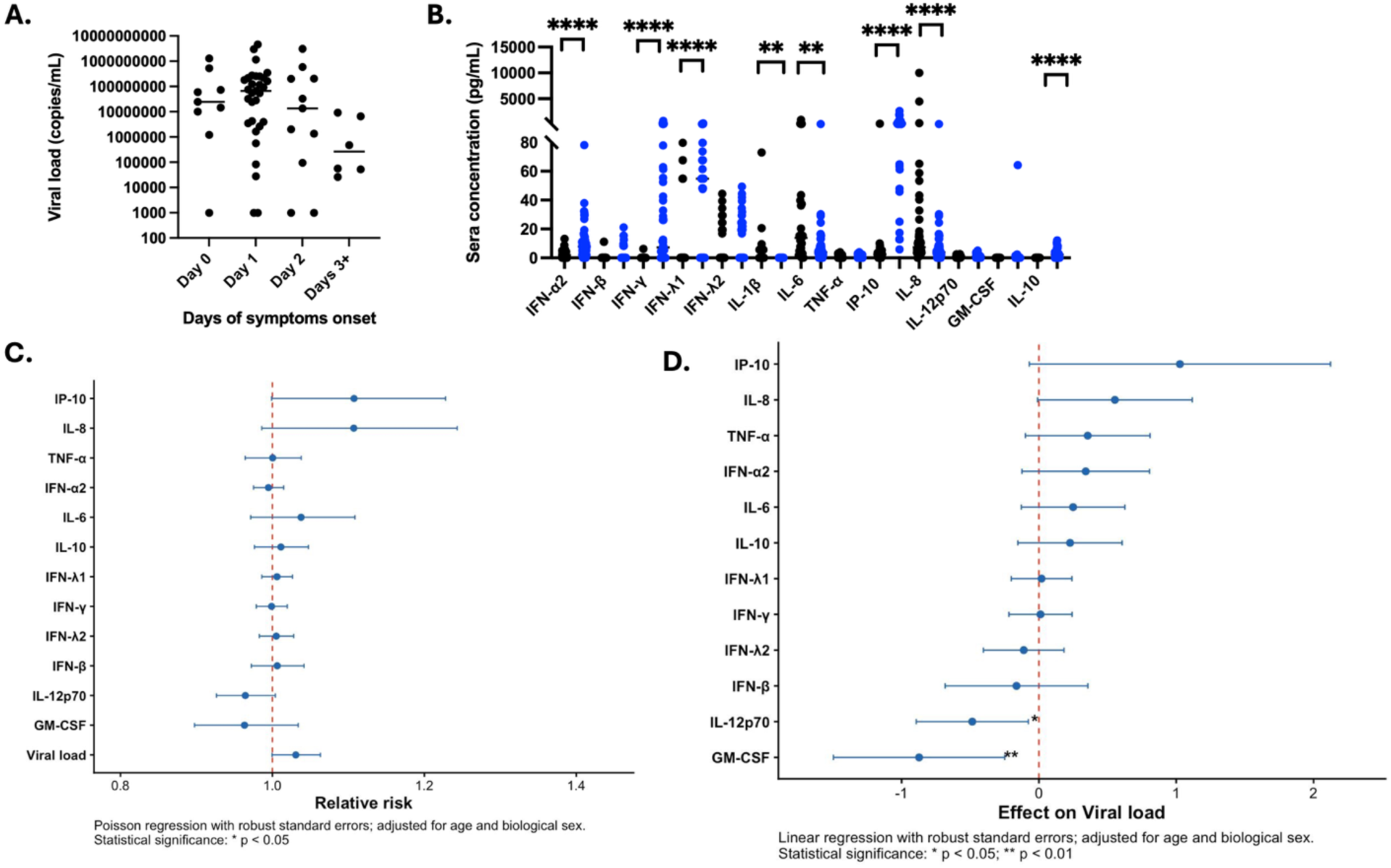
Early OROV acute infection induces a pro-inflammatory anti-viral response. A. Quantification of viral load (copies/mL) in sera of OROV confirmed cases per day of symptom onset. Kruskal-Wallis test, p=0.0797. B. Quantification in pg/mL of pro-inflammatory cytokines (IL-1β, IL-6, TNF-α, IL-12p70) and chemokines (IP-10, IL-8), an anti-inflammatory cytokine (IL-10), a growth factor (GM-CSF), and type 1, 2, and 3 interferons (IFN-α2, IFNγ, IFN-λ1, IFN-λ2) in sera from OROV confirmed cases (blue dot) compared to healthy controls (black dot) from the same region. Mann Whitney test was done for statistical differences between two groups. Statistical significance: *p<0.05, **p<0.01, ***p<0.001, ****p<0.0001. Representation of the RR with its standard deviation obtained through a bivariate regression analysis adjusted for age and sex, to determine the association of each cytokine or factor with early acute infection (2 or less days of symptoms onset) (C) or with viral load (D). Statistical significance: *p<0.05, **p<0.01.

### Clinical characterization of OROV infection and comparative arboviral profiling

Among all OROV-positive participants, the most frequently reported symptoms were headache (71.1%), fever (70.7%), myalgia (43.0%), chills (38.8%), and arthralgia (37.0%), with comparable distributions between men and women. In χ² analyses, fever and rash were strongly associated with OROV infection (*P <* 0.001), whereas myalgia and abdominal pain were also significantly associated (*P* < 0.01), when comparing laboratory-confirmed OROV-positive participants to OROV-negative participants, including both symptomatic individuals who sought care at healthcare centers with suspected arboviral illness and asymptomatic individuals primarily recruited during the community-based field investigation.

Consistent with these findings, bivariable Poisson regression models adjusted for age and sex identified higher frequencies of rash (RR = 1.77; 95% CI: 1.44–2.16), abdominal pain (RR = 1.41; 95% CI: 1.14–1.75), myalgia (RR = 1.21; 95% CI: 1.05–1.39), and retro-orbital pain (RR = 1.17; 95% CI: 1.00–1.37) among OROV-positive participants. In contrast, fever was less frequently observed in this group (RR = 0.77; 95% CI: 0.68–0.89) (Supplementary Table 1).

Using forward stepwise selection based on log-likelihood criteria, the final multivariable model retained rash, fever and myalgia as independently associated symptoms. Rash occurred 67% more frequently among OROV-positive participants (RR = 1.67; 95% CI: 1.36–2.04), and myalgia was 27% more frequent (RR = 1.27; 95% CI: 1.10–1.47), whereas fever was 28% less frequent among OROV-positive individuals (RR = 0.72; 95% CI: 0.62–0.84) (Supplementary Fig. S1).

Comparative analyses revealed distinct clinical features that differentiate OROV infection from other endemic arboviral diseases (Fig. 3). Compared with dengue virus (DENV), OROV infection was associated with increased frequencies of headache (RR = 2.38; 95% CI: 1.74–3.24), arthralgia (RR = 2.32; 95% CI: 1.91–2.80), and diarrhea (RR = 1.94; 95% CI: 1.52–2.47) (Fig. 3A). In contrast to chikungunya virus (CHIKV), OROV infection was characterized by markedly higher frequencies of retro-orbital pain (RR = 3.58; 95% CI: 2.89–4.44) and headache (RR = 2.27; 95% CI: 1.88–2.74) (Fig. 3B). When compared with Zika virus (ZIKV), OROV cases more frequently reported abdominal pain (RR = 1.15; 95% CI: 1.00–1.31) (Fig. 3C). Relative to encephalitic alphaviruses, including Madariaga virus (MADV) and Venezuelan equine encephalitis virus (VEEV), OROV infection was associated with a substantially higher frequency of headache (RR = 2.88; 95% CI: 2.22–3.73) (Fig. 3D).

**Figure 3.**
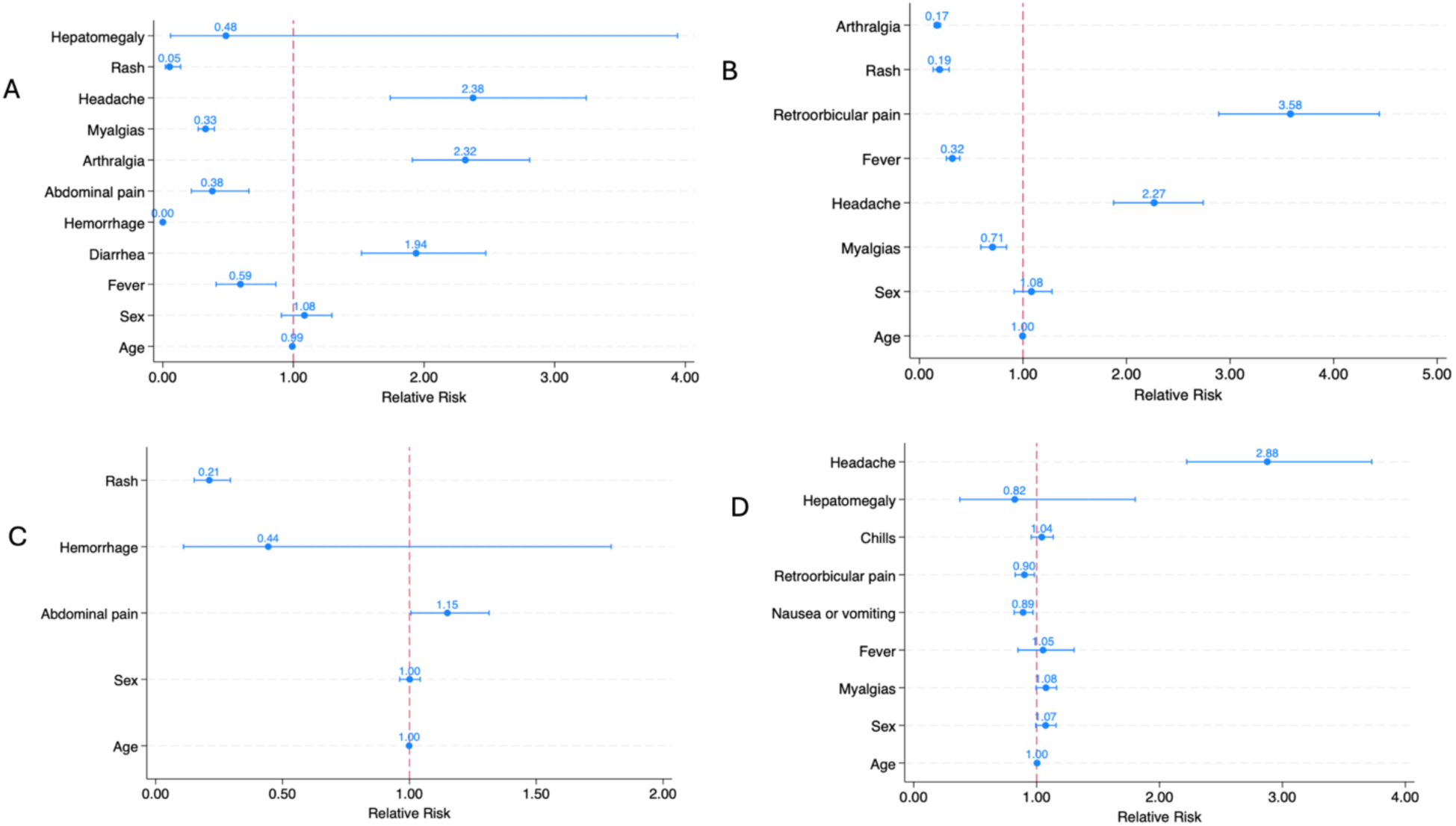
Clinical predictors distinguishing Oropouche virus infection from other arboviral infections. Forest plots showing relative risk (RR) estimates and 95% confidence intervals for clinical variables associated with laboratory-confirmed Oropouche virus infection compared with (A) Dengue virus, (B) Chikungunya virus, (C) Zika virus, and (D) alphaviral encephalitis viruses (Venezuelan equine encephalitis and Madariaga virus). Each panel presents multivariable-adjusted relative risks for demographic and clinical features. The vertical dashed red line indicates RR = 1 (no association). Points represent point estimates and horizontal bars represent 95% confidence intervals. Variables with confidence intervals not crossing 1 are considered statistically significant. All models were adjusted for age and sex.

### Household-level risk factors and transmission dynamics of OROV infection

We implemented a structured household survey to characterize sociodemographic, housing, sanitation, and behavioral determinants of infection among residents of high-incidence communities in Darién. The survey enrolled 289 participants (182 women, 63.0%; 107 men, 37.0%) from 189 households, capturing a wide age range (0–75 years), mixed livelihoods (40.3% homemakers, 27.5% students), and predominantly low-income households (67.0% reporting ≤USD 400/month), according to the poverty and household income distribution report of the Ministry of Economy and Finance (MEF) of Panama. Housing profiles reflected typical rural and peri-urban conditions: 62.8% had concrete floors, 59.4% concrete walls, 55.7% had one or two rooms, and 68.3% relied on flush toilets, while 31.7% used latrines; waste collection services covered 51.1% of households, with the remainder relying on burning or another disposal method (Supplementary Materials, Table S2).

In bivariate analyses, infection risk varied primarily with age and sanitation indicators. The lowest infection rate was reported in the first age quintile (children < 16 years, 67.7%) and fifth age quintile (54-75 years, 67.9%), while working aged adults reported higher rates (Q2 17–27, 82.8%; Q3 28–37 77.2%; Q4 38–55 89.1%). Latrine use was associated with higher infection prevalence compared with flush toilets (86.7% vs. 72.7%; p = 0.009) (Supplementary Material, Table S3).

Multivariable Poisson regression adjusting for age and sex identified a coherent sanitation signal. Residence in households using latrines was associated with an increased risk of infection (RR 1.20; 95% CI 1.07–1.35), although this association attenuated in the fully adjusted model (RR 1.16; 95% CI 0.99–1.35; p = 0.06). This attenuation likely reflects confounding by socioeconomic and community-level factors, as latrine use was more common among lower-income rural households located in high-incidence communities. Together, these findings suggest that the observed association may reflect broader environmental and community-level risk conditions rather than an independent effect of latrine use itself. Household waste burning emerged as an independent predictor of OROV infection (RR 1.18; 95% CI 1.01–1.39; p = 0.037), even after controlling demographic covariates and other housing characteristics.

To further evaluate household-level transmission following viral introduction, we applied a final-size chain binomial framework to estimate the probability of infection following introduction of OROV into a household. Among 44 eligible households, encompassing 76 susceptible individuals and 43 inferred secondary infections, the pooled probability of household infection was 56% (95% CI: 45–68), highlighting a high cumulative risk of infection at the household level once OROV is introduced.

### Community contact tracing

To identify additional infections in the vicinity of confirmed cases, active case finding was conducted among 47 previously identified laboratory-confirmed cases during the surveillance period (December 2024–February 2025), targeting individuals residing in the same household as cases, and nearby households. Among these previously identified cases, 6 (12.8%) were RT-PCR-positive, 8 (17.0%) were IgM-positive, 5 (10.6%) were IgG-positive, and 28 (59.6%) exhibited combined IgM/IgG reactivity at the time of investigation. A total of 242 living in close proximity to the confirmed cases were identified (mean 5.1 contacts per case), with proximity defined in spatial terms (neighbouring or nearby households) rather than direct interpersonal contact. Of these, 175 (72.3%) tested laboratory-positive by RT-PCR and/or ELISA, whereas 67 (27.7%) were negative. Overall, 70 contacts (28.9%) reported at least one symptom (median 7 days, IQR: 3–21), while 172 (71.1%) remained asymptomatic. Among case-contact pairs with available symptom onset dates for both individuals, 87.5% of contacts reported symptom onset after the case, while 12.5% reported symptom onset on the same day. A total of 105/175 laboratory-positive confirmed infections among contacts (60.0%) were asymptomatic.

Among 175 laboratory-positive contacts, 8 (4.6%) were RT-PCR-positive, including 5 symptomatic and 3 asymptomatic individuals, representing the potentially infectious fraction at the time of investigation. IgM seropositivity predominated, detected in 145 contacts (82.9% of laboratory-positive individuals), suggesting that most infections likely represent recent or resolving exposure rather than active infections. Among IgM-positive contacts, 42 reported symptoms between December 2024 and February 2025, whereas most asymptomatic laboratory-positive contacts were identified solely through serological evidence. IgG-only reactivity was identified in 4 contacts (2.3%), and 18 (10.3%) exhibited combined IgM/IgG responses. In the contact tracing component, descriptive information was restricted to contacts (n = 242), excluding cases from this summary. Among contacts, 78.6% reported knowing someone ill outside the household, and 79.0% reported recent contact with a symptomatic individual, most commonly occurring both outside and in the home (35.8%). Travel was frequent, with 45.0% of contacts reporting movement within Darién and 36.7% outside the province. Symptoms occurring within ≤14 days of onset were reported by 65.1% of participants (median 11 days, IQR 3–22.5), most frequently in January (51.0%) and February 2025 (36.9%); 28.9% reported fever at the time of interview. Bivariable comparisons between cases and contacts are presented in (Supplementary Table S4). Details of the bivariable and multivariable regression analyses of cases and contacts are provided in the Supplementary Materials (Table S5).

### Genomic characterization of OROV-positive cases and geographical transmission patterns

We obtained full-length OROV segments from clinical cases identified among permanent residents of Darién Province, Panama, residing in seven townships across Darién Province and the Emberá-Wounaan Comarca (n=34 for L segment, n=34 for M segment, and n=29 for S segment). To investigate whether genetic divergence may have contributed to the delayed recognition of the Panama outbreak relative to widespread transmission elsewhere in South/Central America and the Caribbean, we assessed the molecular characteristics and evolutionary relatedness of locally generated genomes (n=29 after quality curation and segment completeness filtering) against contemporaneous OROV sequences from across the Americas. Phylogenetic analyses incorporated publicly available OROV genomes sampled between 2020 and 2025 from seven countries in the Americas, all with ≥80% segment coverage (n=613 L, n=691 M, and n=647 S; Figure S3, Table S6). This dataset included the most recent pre-outbreak genome from Panama, hOROV/Panama/A003066/2024, detected in August 2024^11^. We reconstructed maximum-likelihood (ML) phylogenies independently for each genomic segment (Figures S4–S6), as well as for concatenated genomes (Figure S7), and evaluated both phylogenetic structure and temporal signal across datasets (Figures S8–S9). ML phylogenies clearly indicated that hOROV/Panama/A003066/2024 did not cluster within or near the monophyletic group of 2024–2025 OROV Panamanian sequences, hereafter designated the OROV_PAN2024-2025_ lineage. Subsequent Bayesian phylodynamic analyses based on the M, L, S, and concatenated segment datasets resolved well-supported maximum clade credibility (MCC) trees, consistently showing that the 2025 Panamanian sequences formed a distinct monophyletic lineage with strong nodal support (posterior probability [PP] ≥ 0.90) (Figure 4; Figure S10). Independent MCC reconstructions of the L, M, and S segments each corroborated the same well-supported monophyletic clustering of OROV_PAN2024-2025_ (PP ≥ 0.90) (Figure S10). L, M and S MCC trees independently corroborated the well-supported (PP≥0.90) monophyletic clades for OROV_PAN2024-2025_ (Figure S10). Relative to contemporaneous regional strains, OROV_PAN2024-2025_ exhibited two notable non-synonymous amino acid substitutions in the M segment, T722N and S1325P, both occurring in coding regions previously implicated in host-virus interactions (Figure S11)^19^. However, branch-specific selection analyses indicated that these substitutions are evolving under neutral to weakly diversifying selection, suggesting they are unlikely to confer functional advantages related to transmission efficiency or virulence (Table S7). The absence of positive selection at T722N and S1325P indicates these substitutions arose through genetic drift rather than adaptive evolution, consistent with recent introduction rather than gradual adaptation during cryptic persistence.

**Figure 4.**
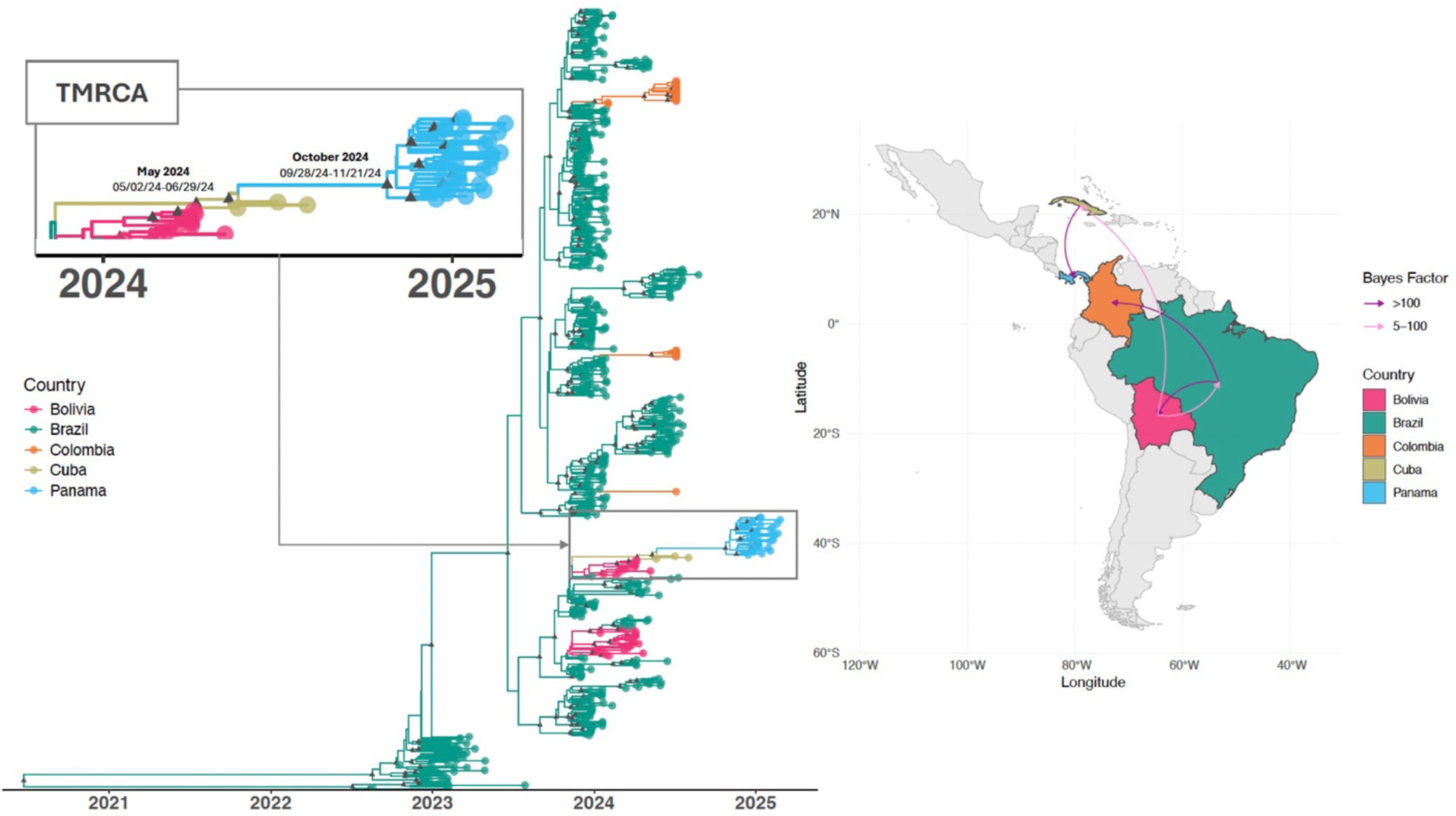
Spatial dissemination of Oropouche virus from 800 full-genome sequenced samples from 2020 to 2025 confirmed cases in Latin America. (a) Bayesian scale MCC tree under a discrete phylogeographic model after concatenating the three genomic segments (L, M, and S). Branches and tips are colored according to the most probable ancestral location, with a posterior probability (PP) ≥0.90 (black triangle). (b) The boxplot shows the evolutionary rate with the molecular clock median value and 95% HPD. (c) The map shows the spatial spread of OROV among eight sampled locations, with transition routes in the region supported by Bayes Factor (BF) evidence (very strong support: BF ≥100; strong support 5 ≤ BF < 100).

Discrete phylogeographic reconstruction revealed multiple well-supported dispersal routes at both geographic scales, across the Americas and within Panama (Table S8). The origin of OROV_PAN2024-2025_ in Panama, inferred from molecular dating of the concatenated dataset, was strongly supported by a viral movement event linking Cuba to Panama (BF = 324.4) (Figure 4, Table S8). The time to the most recent common ancestor (TMRCA) for this introduction was estimated to be October 2024 (95% highest posterior density [HPD]: September–November 2024). Comparable estimates were recovered across individual genomic segments, with TMRCAs of October 2024 for L (95% HPD: September 2024–February 2025), November 2024 for M (95% HPD: October–December 2024), and December 2024 for S (95% HPD: November–December 2024). Earlier migration events were inferred from Brazil to Bolivia and Colombia with strong support (BF ≥ 100), and from Bolivia to Cuba and Bolivia to Brazil with moderate support (5 ≤ BF < 100) (Table S8). The median estimated evolutionary rate was 1.6×10^−3^ substitutions/site/year for the concatenated genome dataset, consistent with segment-specific estimates of 1.6×10^−3^, 1.6×10^−3^, and 5.8×10^−3^ for the L, M and S segments. Greater uncertainty was observed for the shorter S segment, consistent with previous studies^12^ (Figure S12).

To further characterize local transmission dynamics within Panama, we implemented an asymmetric diffusion model and reconstructed spread across eight locations (Cémaco, El Real, Lajas Blanca, Metetí, Río Iglesias, Sambú, Yape and Yaviza) using 29 genomes. Most sequences clustered within a monophyletic clade circulating between the Darién and Emberá provinces. The analysis revealed concurrent dispersal of two well-supported Panamanian lineages moving from Metetí to Yaviza (both corregimientos in Darién province) with strong statistical support (BF > 100). This two-lineage pattern was also recovered when genomic segments were analysed independently (Figure S12). Additional diffusion routes to Cémaco, El Real, Lajas Blanca, Río Iglesia, Sambú, and Yape were supported with moderate evidence (5 ≤ BF < 100) (Figure 5; Table S8). These findings suggest dual seeding events that established distinct transmission chains among geographically connected communities, consistent with known mobility and incidence hotspots in the region (Table S8). The TMRCA for these two Panamanian lineages, estimated from the concatenated dataset, was November 2024 (95% HPD: July–December 2024). Similar estimates were obtained across genomic segments: September 2024 for L (95% HPD: June–November 2024), October 2024 for M (95% HPD: October–November 2024), and November 2024 for S (95% HPD: September–November 2024). The median evolutionary rate for the national dataset was 1.8×10^−3^ substitutions/site/year, with broadly comparable segment-specific estimates of 8.9×10^−4^, to 1.6×10^−3^, and 1.0×10^−3^ substitutions/site/year for L, M, and S, respectively (Figure S12). As above, uncertainty was greatest for the shorter S segment. Taken together, the national-and regional-scale phylogeographic analyses indicate that the Panamanian outbreak likely emerged between June 2024 and December 2024 (Figures 4 and 5).

**Figure 5.**
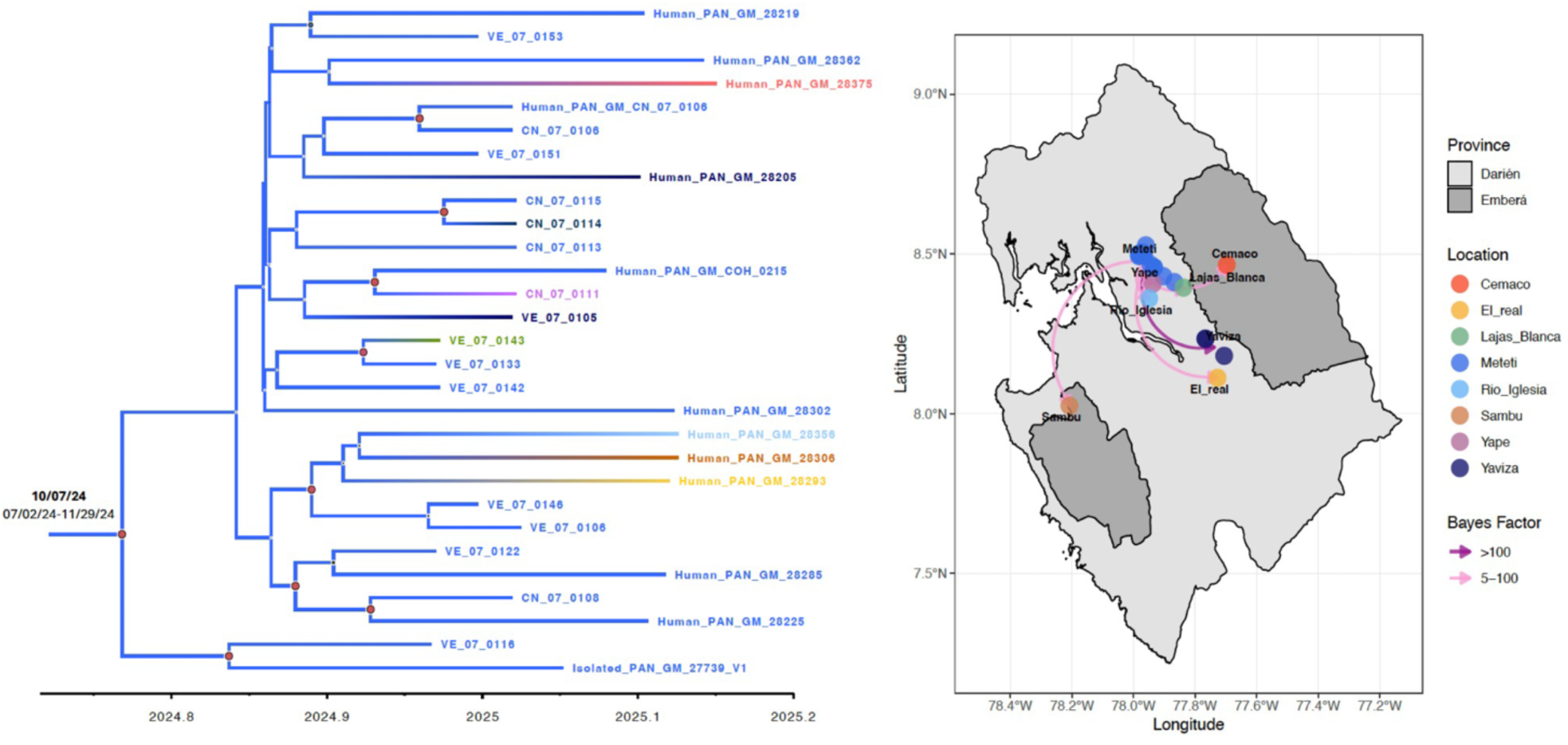
Spatial dissemination of OROV_PAN2024-2025_ clade across the Darién and Emberá provinces, Panama. Bayesian scale MCC tree under a discrete phylogeographic model after concatenating the three genomic segments (L, M, and S) (n=29 full-length genomes). Branches and tips are colored according to the most probable ancestral location, with a posterior probability (PP)≥0.90. The accompanying map shows the spatial spread of OROV among eight sampled locations, with transition routes supported by Bayes Factor (BF) evidence (very strong support: BF ≥100; strong support 5 ≤ BF < 100).

### Risk of OROV outbreak following introduction

Using surveillance data collected in Brazil during the early stages of Oropouche expansion (July 2021–early 2025), we created a model that identified specific weather, environmental, agricultural, and human predictors of OROV outbreak risk. The sources and original units of all predictors are listed in Table S10. Predictors were z-score normalized prior to model fitting. For most predictors, the range of values within Panama fell within the range seen in Brazil, and no values substantially exceed the range seen in Brazil; see Figure S13 for ranges of predictors.

Outbreak risk was positively associated with average daily vapor pressure (a measure of humidity) in the 30 days preceding an initial case report (β = 1.20, p < 0.001), with average daily precipitation (β = 0.583, p < 0.001) and the average daily high temperature (β = 0.972, p < 0.001) in the 30 days following an initial case report, and with the amount of precipitation typically received in the driest month (β = 0.262, p = 0.008), but negatively associated with the average daily low temperature in the 30 days preceding an initial case report (β = –1.13, p = 0.011) and with the amount of precipitation typically received in the wettest month (β = –0.511, p < 0.001). The percent of land dedicated to coffee cultivation (β = 0.234, p = 0.002) and to the cultivation of tropical and subtropical fruits (β = 0.263, p < 0.001) were associated with increased risk of outbreak, while the percent of land dedicated to cereal crops was negatively associated with outbreak risk (β = –1.39, p < 0.001). Finally, the maximum human footprint value within the municipality (the maximum value of a measure of human development) was positively associated with outbreak risk (β = 0.468, p < 0.001), while the log_10_ density-weighted population density (the population density most people experience) was negatively associated (β = –0.48, p = 0.00001). Further details on the predictors, model development, and final estimates can be found in Tables S10 through S13.

### Mapping Outbreak Risk Within Panama

We projected risk at the corregimiento (sub-district; roughly equivalent to a county) level using the model obtained from the fit to data from Brazil. This prospectively identified Darién/Emberá as highest risk (mean 71–73%); Pinogana (1,162/100k incidence) ranked top decile, consistent with nationwide projections (Figure 6A). Projections of outbreak risk within Panama using climate normals (“typical” monthly weather derived from long-term climate data)^17^ predict overall high risk of outbreak following introduction (mean of corregimiento-level risk = 56%). There is substantial geographic variation in risk (Figure 6A); the 10th percentile of corregimientos have a mean estimated outbreak risk of 28.4%, while the 90th percentile of corregimientos have a mean outbreak risk of 76.14%. The provinces with the highest risk included the Darién province (mean risk over “typical” year and corregimientos = 71.2%) and the adjacent region of Emberá (73.1%), as well as Bocas del Toro (mean risk over “typical” year and corregimientos = 73.9%) and Los Santos (mean risk over “typical” year and corregimientos = 72.7%). In contrast, the highly populous Panamá province has far lower average risk (43.3%), though it contains high-risk districts (Chepo; mean risk over “typical” year and corregimientos = 63.44%, and Chimán; mean risk over “typical” year and corregimientos = 66.2%,) that are adjacent to Darién province. This is due to the negative association between population density and outbreak risk; see Figure 6D for geographic distribution of human density within Panama.

**Figure 6.**
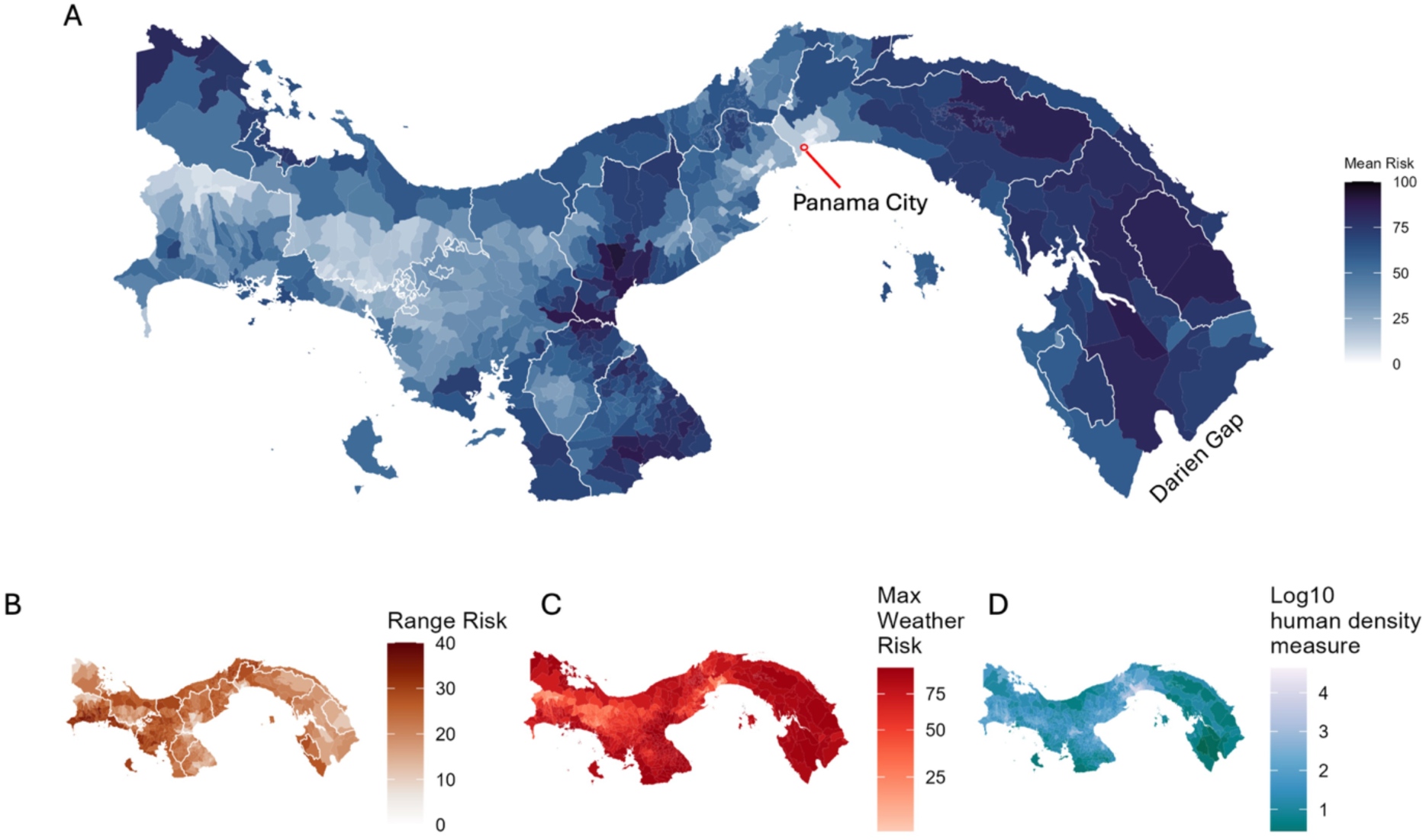
Mapping risk of outbreak at the corregimiento level, based on the model fit to Brazilian OROV introduction outcomes. **A)** Risk estimated using monthly climate normals, averaged over all months. Regions outlined in white. B) The difference (range) between highest and lowest risk month estimated from climate normals for each corregimiento, C) Risk in highest-risk month for each corregimiento estimated from weather conditions 2023–2025, representing the most favorable weather conditions for transmission observed during that period, D) Corregimiento-level log_10_ density-weighted population density (the average population density people experience), a negative correlate of outbreak risk.

Risk varies modestly throughout the year; the average difference at the corregimiento level between months with maximum and minimum risk estimated from climate normals is 25.6% (Figure 6B). While climate normals allow us to forecast general patterns in seasonality (Figure S16, Figure S17), weather often deviates from long-run averages, as do our risk estimates. Notably, in the years 2023–2025, weather in November and December resulted in higher risk estimates than would be expected from long-run averages across much of the country (Figure S18). Additionally, areas with typically low or moderate risk may transiently experience elevated risk due to weather conditions. Figure 6C shows the highest risk estimate from weather in the period 2023–2025 for each corregimiento. Notably, in the years 2023–2025, weather in November and December resulted in higher risk estimates than expected from long-run averages across much of the country (Figure S18). This anomalous elevation coincides with the phylogeographic TMRCA (Oct 2024; 95% HPD Jul–Nov).

## Discussion

Our findings resolve the Darien paradox of OROV emergence nearly one year after Brazil’s 2024 surge. Integrated surveillance reveals a single recent introduction in October 2024 from a Cuba-linked ancestor (Bayes factor 324.4), igniting an outbreak rather than prolonged cryptic circulation, precisely when risk maps show Darien weather peaking above climate normals. Local diversification produced two lineages spreading from Meteti to Yaviza along high-mobility corridors (BF >100), matching incidence hotspots in Pinogana district (1,162 per 100,000). Neutral M-segment evolution (T722N, S1325P substitutions under purifying selection) argues against adaptive escape, aligning with regional patterns of the reassortant OROV_BR-2015-2024_ lineage’s ecological expansion rather than viral adaptation^12,20^.

OROV spreads efficiently within households. The 56% secondary attack probability exceeds typical dengue estimates of 30 to 40% and approaches values reported for chikungunya in immunologically naive populations. In the largest prospective contact-tracing study of OROV to date, 60% of infected contacts were asymptomatic or minimally symptomatic. This subclinical reservoir means containment cannot rely on clinical case-finding alone.

At the community scale, Darién’s rural sanitation deficits, low population density (risk-amplifying per model), and extreme mobility (500,000 annual crossings) converted a single seeding into 752 per 100,000 incidence with focal intensity. Waste burning remained independently associated with OROV infection in the multivariable model (RR = 1.18). Although entomological data were not available in the present study, previous studies have suggested that environments with accumulated organic waste and humid decomposing substrates may favor peridomestic proliferation of *Culicoides paraensis* and other potential OROV vectors^21^. Therefore, this association may reflect increased exposure to biting midges in settings where household waste accumulates prior to burning. Because waste burning is a potentially modifiable practice, improving community-level waste management and collection systems could represent a feasible intervention to reduce peridomestic vector exposure. Coffee and tropical-fruit agriculture also predicted higher outbreak probability, contrasting with urban *Aedes*-driven dengue, Zika, and chikungunya transmission, and consistent with peridomestic midge exposure where avoidance is impossible^15,22^. Twenty-nine infections (6.5%) occurred among non-resident personnel (e.g., border service) returning to other provinces, providing direct epidemiological evidence of mobility-driven dispersal beyond the outbreak’s focus^13,23^.

OROV evaded routine surveillance for six weeks not because of technical limitations, but because of structural diagnostic failure. Despite 43% positivity among evaluated individuals, OROV mimics dengue clinically yet shows a distinctive syndromic signature: higher frequencies of headache and retro-orbital pain than dengue or chikungunya (Fig. 3), and an early cytokine profile dominated by 80-fold IP-10 elevation within 48 hours alongside coordinated type I, II, and III interferon responses. This molecular window opens precisely when RT-PCR sensitivity wanes after day 5, so paired syndromic and immunologic signatures can guide targeted confirmation where febrile illness is reflexively attributed to dengue. The paradoxically lower fever frequency among OROV-positive cases (multivariable RR 0.72) may reflect surveillance bias, as febrile patients are preferentially tested for dengue. Combined with the short RT-PCR window and the high proportion of asymptomatic infections identified during community-based investigation (53.8%), these findings indicates that passive, symptom-triggered surveillance substantially underestimates OROV burden, creating a blind spot that will likely recur across dengue-endemic Central and South America unless febrile algorithms explicitly include OROV^24^. Manaus blood donor data, with an estimated 390,000 infections during Brazil’s 2023 to 2024 outbreak^25^, confirm that fever-based case definitions miss most transmission and that serology-based surveillance is needed to capture community-level circulation.

Models trained on Brazilian surveillance data (July 2021 to early 2025) prospectively identified Darién and Emberá as highest risk. Forecasts indicate broad Panamanian vulnerability, with a mean predicted outbreak probability of 56% at the corregimiento level following introduction; Bocas del Toro (74%) and Los Santos (73%) emerged as the next highest-risk provinces, neither with reported OROV cases. If outbreaks emerge there during 2025 to 2026, this would validate the model’s prospective utility; sustained absence would instead indicate additional demographic or surveillance constraints, informing model refinement. The pattern is not uniformly permissive: high-density Panamá province registered a lower mean probability (43%) despite containing high-risk districts such as Chepo and Chimán, showing that demographic structure modulates risk beyond climate. Anomalous late-year weather across 2023, 2024, and 2025 elevated risk above climate normals, aligning temporally with the phylogeographic TMRCA of October 2024 (95% HPD July to November). Beyond prior Brazilian genomic^12,14^ and ecological^15^ work, this is the first integrated Central American analysis to resolve emergence timing, validate cross-country risk prediction, and link peridomestic exposures to transmission dynamics.

Panama’s position at the South-Central America interface, with Darién Gap migration and Panama Canal traffic, makes it a continental bottleneck for arboviral dissemination. Three immediate changes are warranted. First, deploy IgM ELISA alongside RT-PCR at primary healthcare facilities in Darién and adjacent provinces. Since 95% of asymptomatic infections were detected by serology rather than molecular testing, earlier deployment would have shortened the six-week detection delay to under two weeks. Second, extend sentinel genomic surveillance to Bocas del Toro and Los Santos before the predicted November to December 2025 risk peak, prospectively testing model forecasts. Third, integrate OROV into routine arboviral panels for undifferentiated febrile illness. This requires policy change, not only technical capacity: regional health ministries should expand febrile algorithms to include OROV RT-PCR and ELISA as reflex tests alongside dengue. Weather-informed early warnings can further guide preemptive resource allocation, preventing OROV from becoming entrenched as dengue has. These principles generalize to other cryptic arboviruses in dengue-endemic settings, where structural diagnostic biases systematically delay recognition of novel threats.

Several limitations warrant consideration. Brazilian risk models may not generalize to Panama despite strong predictive performance (e.g., high predicted risk in Darién and Chepo hotspots). Projections rely on climate normals and 2023 to 2025 weather but cannot anticipate unmeasured extremes altering *C. paraensis* population dynamics^22^. We lack entomological data; vector density, distribution, and biting rates remain uncharacterized. Risk factor associations may reflect residual confounding, and household transmission models assume uniform mixing, given no evidence the vector exhibits age-specific feeding preferences. Genomic sampling from seven townships is robust for phylogeography but could miss cryptic diversity from unsampled communities or earlier introductions. Future work should: (i) couple direct *C. paraensis* surveys with household-level infection data in high-versus low-risk communities; (ii) test the model prospectively as 2025 to 2026 unfolds; and (iii) stratify behavioral risk beyond current environmental proxies.

Darién is a sentinel for OROV’s northward trajectory, and a template for finding the next cryptic arbovirus before it spreads.

## Materials and Methods

### Ethics statement

All human participant research, including febrile illness, migrant, community surveillance, and contact tracing activities, was conducted in accordance with the Declaration of Helsinki and national regulations of Panama. Study protocols were reviewed and approved by the Comité de Bioética de la Investigación del Instituto Conmemorativo Gorgas de Estudios de la Salud (CBI–ICGES). Approved projects included: Oropouche surveillance (FID2024-102; No. 223/CBI/ICGES/24), SIVAT surveillance platform (FID2023-147; No. 274/CBI/ICGES/23), longitudinal cohort study (FID2021-96; No. 138/CBI/ICGES/22), febrile illness surveillance (FID-146; No. 281/CBI/ICGES/23), and migrant health study (FID2024-101; No. 028/CBI/ICGES/26). Written informed consent was obtained from all adult participants and from parents or legal guardians of minors prior to enrollment.

### Outbreak detection and One Health surveillance in Darién

Outbreak detection was enabled through an integrated One Health surveillance and cohort initiative, called the Darién Gap Cohort, implemented by the Carson Centre for Research in Health and Ecosystems in Darién Province, Panama. This platform integrates longitudinal surveillance of farmers and Indigenous communities, livestock and wildlife monitoring, and syndromic febrile surveillance across regional health centers^26^. The first reports of unusual febrile illness were noted on 18 December 2024, from a participant enrolled in the Darién Gap Cohort. Subsequently, on 20 December 2024, clinicians at the Metetí Health Center in Darién independently reported an apparent increase in febrile disease cases. In response, clinical samples were submitted for diagnostic testing targeting common endemic pathogens, including dengue, malaria, rickettsiae, leptospirae, and the encephalitic alphaviruses Venezuelan equine encephalitis virus (VEEV) and Madariaga virus (MADV); all results were negative. In parallel, a multiplex global febrile illness panel (FilmArray® Global Fever Panel – RUO) screening for 19 pathogens was applied, with no pathogens detected^27^. On 13 January 2025, a multidisciplinary clinical and field team from the Carson Centre conducted active surveillance in the community of Metetí, collecting additional clinical samples and evaluating additional febrile cases. These samples were screened for endemic arboviruses, and, in this round of investigation, testing was expanded to include RT-PCR for OROV and viral isolation. On 25 January 2025, circulation of OROV in Darién was detected by Carson center and reported to MINSA, then confirmed by ICGES-Panama.

### Oropouche Case Definition

We used the case definitions for OROV established by the Pan American Health Organization^28^. A suspected Oropouche case was defined as any person that presented fever (T ≥38°C) acutely with at least one of the following symptoms: headache, chills, nausea, or vomiting, and a probable case as a suspected case that has an epidemiological link to a confirmed case of Oropouche. We defined a confirmed case as a probable or suspected case that had laboratory confirmation either through viral isolation, RT-PCR, or IgM ELISA (with seroconversion of antibodies or a fourfold or greater increase in the concentration of antibody titers of paired samples taken more than 7-10 days apart) (Figure S2).

### Contact tracing

Between January and February 2025, a follow-up investigation of OROV-confirmed cases and their contacts was conducted in multiple communities across Darién province. Contact tracing was initiated immediately after laboratory confirmation by the Carson Centre and the regional laboratory of the Gorgas Memorial Institute in Metetí. Contacts were defined as individuals residing in the same household as a previously confirmed case or in adjacent homes. Active case findings were conducted in all visited households to identify any individuals with current or recent febrile illness, regardless of contact status. Three field teams operated simultaneously to optimize coverage. Upon informed consent, participants were enrolled and interviewed using a standardized REDCap-based questionnaire that assessed sociodemographic factors, housing conditions, recent travel, exposure history, and symptoms (Questionnaires included as supplementary appendix). Individuals previously diagnosed with OROV were additionally evaluated for persistent symptoms and potential post-viral sequelae.

### Sample collection and processing

Biological samples were collected from all participants. A 10 mL venous blood sample was obtained from each contact individual, as well as from any newly identified individual presenting with acute febrile illness at the time of the visit. In addition, a urine sample was collected from each participant for diagnostic purposes. All specimens were stored and transported under appropriate biosafety conditions and cold chain protocols to ensure the integrity of samples for subsequent molecular and serological analyses.

## Laboratory Testing

### Viral isolation and RT-PCR detection of OROV

Serum samples obtained during the acute stage of disease (duration of symptoms, ≤5 days), was used to inoculate monolayers of African green monkey kidney epithelial cells (Vero E6) and observed for cytopathic effects. RNA from clinical samples and isolates was extracted using an automated KingFisher Flex purification system (Thermo Fisher Scientific) with the MagMAX Viral/Pathogen Nucleic Acid Isolation kit, following the manufacturer’s instructions. RT-PCR was performed using the iTaq Universal Probes One-Step Kit (Bio-Rad, USA), employing primers and probes specific for OROV (Spplementary Materials, Table S9), previously validated^29^. Amplification was carried out over 45 cycles, and samples with a cycle threshold (Ct) value ≤40 were considered positive for OROV RNA.

### Serological testing

Serological analyses included ELISA and plaque reduction neutralization testing (PRNT). Anti-OROV IgM and IgG antibodies were detected using recombinant protein-based ELISA kits (Euroimmun, Germany)^30^. PRNT was performed using heat-inactivated sera incubated with 1,000 PFU/mL of the OROV strain TVP6135, and neutralizing antibody titers were defined as the highest serum dilution achieving ≥90% plaque reduction (PRNT₉₀).

### OROV viral load quantification by real-time RT-PCR

OROV RNA loads were quantified using a plasmid containing the complete S segment (VectorBuilder ID: VB250815-1028ady; pRP-Amp-OROV_S_951 bp) as an absolute quantitative standard. Plasmid concentration was calculated based on molecular weight, and a 10-point, 10-fold serial dilution (10⁰–10¹⁰ copies/µL) was prepared in nuclease-free water and included in duplicate in each RT–qPCR run.

Total viral nucleic acids were extracted from 200 µL of serum using the MagMAX Viral/Pathogen Nucleic Acid Isolation Kit on a KingFisher™ automated platform (Thermo Fisher Scientific), following the manufacturer’s instructions, with elution in 60 µL nuclease-free water. Extracted RNA was stored at −80 °C until analysis.

OROV RNA detection and quantification were performed using a one-step RT–qPCR assay as previously described^28^, using the SuperScript™ III Platinum® One-Step qRT-PCR Kit (Thermo Fisher Scientific). Reactions were performed in a final volume of 25 µL containing 5 µL of extracted nucleic acid and run on a QuantStudio™ Real-Time PCR System under the following cycling conditions: 52 °C for 15 min, 94 °C for 2 min, followed by 45 cycles of 94 °C for 15 s, 55°C for 40 s, and 68 °C for 20 s. The fluorescence threshold was manually set; samples with Ct ≤40 were considered positive. Each run included no-template controls, a negative clinical control, and the full plasmid standard curve. Viral RNA loads were calculated from the standard curve and expressed as log₁₀ copies/mL.

### Cytokine profiling

OROV-RT-PCR positive samples from participants with ≥ 15 years old and with ≤ 7 days of symptoms onset were selected, as well as healthy (asymptomatic) OROV-negative individuals from the same region, matched for age-range and sex, to analyze early immune responses and their association to clinical symptoms. Host immune responses were assessed using the LEGENDplex™ Human Anti-Virus Response Panel V02 (13-plex; BioLegend), a multiplex bead-based immunoassay that quantifies interferons (IFN-α2, IFN-β, IFN-γ, IFN-λ1/IL-29, IFN-λ2/IL-28A), pro-inflammatory cytokines (IL-1β, IL-6, TNF-α, IL-12p70), chemokines (IP-10/CXCL10, IL-8/CXCL8), GM-CSF, and IL-10. Assays were performed according to the manufacturer’s protocol. Briefly, samples were incubated with fluorescent capture beads, followed by biotinylated detection antibodies and streptavidin–phycoerythrin. Cytokines were quantified using seven-point standard curves. Data acquisition was performed on a BD FACSFortessa™ X20 flow cytometer. Data were analyzed using LEGENDplex™ Data Analysis Software (v8.0). Cytokine concentrations were expressed in pg/mL.

For statistical comparison, differences between healthy controls and OROV acute patients associated to categorical values (ej. Sex) a Chi.square test was used, whereas for age and other continuous values Mann Whitney test was done. For comparison between different categories, a Kruskal-Wallis test was employed.

A bivariate regression analysis was performed to evaluate the association between immunological and virological markers with different outcomes, consistently adjusting for age and sex. First, early onset of symptoms (defined as ≤ 2 days versus > 2 days from onset) was analyzed as a dichotomous outcome using Poisson regression models with log link and robust variances, which allowed us to estimate relative risks (RR) directly, avoiding the overestimation associated with logistic models in frequent outcomes. Second, viral load was analyzed as a continuous outcome using linear regression models, given its approximately normal behavior after logarithmic transformation. In both approaches, each predictor (cytokines and viral load) was evaluated individually in bivariate models adjusted for age and sex. All continuous predictor variables were analyzed on a natural logarithmic scale to reduce the asymmetry of their distributions, mitigate the influence of extreme values, and facilitate a proportional interpretation of the effects. The results of the regressions were summarized using forest plots, which present the point estimates and their 95% confidence intervals.

### Household risk factors and clinical phenotype modelling

The primary outcome was OROV infection, defined as the PCR-confirmed presence of viral RNA or anti-OROV IgM seropositivity by ELISA and treated as a binary variable (0 = negative, 1 = positive). We adopted a two-stage analytical strategy focused on laboratory-confirmed OROV infections. First, we modelled risk factors associated with laboratory-confirmed OROV infection. Independent variables included sociodemographic characteristics, environmental and behavioral exposure factors, and selected clinical indicators. Descriptive analyzes were performed using contingency tables to summarize frequencies and proportions. Bivariate associations were assessed using Pearson’s chi-square or Fisher’s exact tests, as appropriate. Variables were selected for multivariable modelling based on epidemiological plausibility and prior evidence. Multivariable generalized linear models (GLM) with a Poisson distribution and robust variance were constructed to estimate adjusted relative risk (aRRs) and 95% confidence intervals, identifying independent predictors of OROV infection.

Second, we modelled comparative clinical symptom profiles to characterize the phenotypic presentation of laboratory-confirmed OROV relative to other arboviral infections, including dengue, Zika, chikungunya, and alphavirus-associated encephalitis. Regression-based analyzes were conducted using generalized linear models (GLM) with a Poisson distribution and robust variance to identify distinguishing and overlapping symptom patterns across viral etiologies. Data were analyzed using Stata version 18 (StataCorp, College Station, TX, USA).

Households were sampled from communities with intense OROV transmission identified through surveillance, and the questionnaire (in Supplementary materials) captured individual sociodemographic characteristics, household structure, water and sanitation infrastructure, waste management, and behaviors relevant to vector exposure (for example, time spent outdoors and agricultural activities). Details on the generalized linear models (GLMs) to estimate adjusted aRRs shown in Supplementary Methods.

Additionally, we quantified within-household transmission using a chain binomial final-size mode^31^. Details on determination of household membership, individuals’ infection status, and model structure are in Supplementary Methods.

### OROV genome sequencing

Total RNA extractions and library construction were prepared for shotgun sequencing, using Illumina NovaSeq 6000 platform using S4 and SP flow cells (See Supplementary Methods).

Raw sequencing reads generated were quality-filtered and trimmed. Trimmed reads were processed for taxonomic classification and de novo assembly. Sequence identification, protein comparisons and visualization were performed for taxonomic visualization and assessment. Read mapping was performed to validate organism detection and estimate genomic coverage. De novo assembled OROV contigs were extracted and imported to identify and translate open reading frames, followed by pairwise sequence comparisons to assess percent identity and amino acid differences (See Supplementary Methods).

### Molecular signature, recombination, phylogenetics and phylogeographic analysis

Full genome sequencing of 800 available OROV strains with specified geographical location, time of isolation, and source were downloaded from GenBank (Table S4). Recombinant regions were removed using RPD5 for each OROV viral RNA segment; classified as small (S), medium (M), and large (L). Branches under positive selection were identified, using the hyphy package (https://www.datamonkey.org/)^32–34^. Signature pattern analysis was performed using VESPA^35^. Maximum likelihood inference was carried out using IQTree v2.4.0. Phylogeographic reconstruction with diffusion in discrete space was implemented in BEAST v10.5.0 (See Supplementary Methods).

### Creation of Outbreak Risk Model and Projection in Panama

The risk model was developed using Oropouche case data obtained from the Brazilian Ministry of Health. In short, cases were grouped by municipality and time period; three or more cases separated by no more than 21 days was considered an outbreak, while those that did not meet these criteria were classified as isolated cases. For each municipality, we acquired data on time-invariant features including human density, typical climate, land use for specific crops and types of crops, human footprint, soil conditions, as well as time-varying weather conditions surrounding the initial case of each introduction event. We then developed a generalized linear modeling (GLM) framework to identify features and weather conditions of municipalities where introduction resulted in (i) outbreak or (ii) no outbreak, and to quantify their effects on outbreak risk (detailed in Table S13). See Supplementary Methods for details of data acquisition and model fitting processes.

To create risk maps for Panama, we supplied corregimiento-level predictors and the model described above to the “predict” function from the ‘stats’ package. Risk was estimated using both long-run climate normals as a proxy for typical weather conditions and using observed weather conditions for years 2023–2025. See Supplementary Methods for details on data sources and model fitting.

## Data Availability

All data produced in the present study are available from the authors upon reasonable request and appropriate justification. Some datasets were generated and are maintained by the research group, whereas other data belong to the Ministry of Health of Panama and may be subject to institutional restrictions and data-sharing agreements.

## Acknowledgments

We thank the Ministry of Health (MINSA) for facilitating connections with local health centers and the National Secretariat of Science, Technology, and Innovation (SENACYT) for funding the primary study. AM, JMP, SLV are members of the Sistema Nacional de Investigación (SNI AIP), Panamá, Panamá

## Contributions

Conceived and designed the study: AIB, CM, and JPC

Survey instrument design: AC, XR, JFG, LFR, JPC.

Collected data: WMS, AC, XR, JFG, CLC, YJ, LFR, KEL, AAS, AGA, SCD, JA, BR, JPC, SLV, DB, VS, HC, CJ.

Performed laboratory analysis: XR, CLC, JG, YJ, DB, DRS, MK, DWA, RC, IAG, AIB, CM, JPC, LWA, JGPJ, SLV, YT, SA. JMP, FR, EA. MCG, BM, AYV, ZGR, JA, EWV, NMP.

Medical examinations and/or field investigations: XR, SLV, DB, RC, XR, JFG, CLC, YJ, LFR, KEL, AAS, AGA, SCD, JA, BR, JPC, VS, HC, CJ, JMP, MEA, AYV, ZGR, JA, EWV, NMP,

Performed sequencing and/or sequence validation: LTR, GEV, QKT, ACIV, MF, AL, LTCS, RZC, FM, OC, JG, AM, CA, AM.

Formal phylogenetic analysis: BG, CM, JGPJ, AM.

Formal statistical and modeling analysis: XR, JPC, CAD, ILD; MK, NRF, AIB, LWA

Project implementation: AIB, CM, JPC, LWA, JGPJ

Funding acquisition: AIB, CM, AMYV, MK, MLN, DRS, KH, YT, SA, SLV, NV, and JPC

Wrote the manuscript: AIB, CM, SLV, JPC, LWA, JGPJ

Edited the manuscript: all authors.

All authors read and approved the contents of the manuscript.

## Corresponding authors

Address correspondence to Jean-Paul Carrera, Carla Mavian, Ana I. Bento, Kimberly A. Bishop-Lilly.

## References

1. Pan American Health Organization. Public Health Risk Assessment related to Oropouche Virus (OROV) in the Region of the Americas – 3 August 2024. https://www.paho.org/en/documents/public-health-risk-assessment-related-oropouche-virus-orov-region-americas-3-august-2024 (2024).

2. Braga, A. et al. Oropouche virus infection in pregnancy: emerging evidence on vertical transmission and perinatal outcomes. J. Matern. Fetal Neonatal Med. 39, 2603781 (2026).

3. Castilletti, C. et al. Replication-Competent Oropouche Virus in Semen of Traveler Returning to Italy from Cuba, 2024. Emerg. Infect. Dis. 30, (2024).

4. De Souza, W. M. et al. ICTV Virus Taxonomy Profile: Peribunyaviridae 2024: This article is part of the ICTV Virus Taxonomy Profiles collection. J. Gen. Virol. 105, (2024).

5. Travassos Da Rosa, J. F., et al. Oropouche Virus: Clinical, Epidemiological, and Molecular Aspects of a Neglected Orthobunyavirus. Am. Soc. Trop. Med. Hyg. 96, 1019–1030 (2017).

6. Hughes, H. R. et al. ICTV Virus Taxonomy Profile: Peribunyaviridae. J. Gen. Virol. 101, 1–2 (2020).

7. Gallichotte, E. N., Ebel, G. & Carlson, C. J. Vector competence for Oropouche virus: a systematic review of pre-2024 experiments. Preprint at 10.1101/2024.10.17.24315699 (2024).

8. Cain, M. & Ly, H. Oropouche virus: Understanding “sloth fever” disease dynamics and novel intervention strategies against this emerging neglected tropical disease. Virulence 15, 2439521 (2024).

9. Godinho, I. P. et al. Insights into the expansion of Oropouche virus in Brazil: epidemiological and environmental aspects. Exp. Biol. Med. 250, 10647 (2025).

10. Tesh, R. B. The Emerging Epidemiology of Venezuelan Hemorrhagic Fever and Oropouche Fever in Tropical South Americaa. Ann. N. Y. Acad. Sci. 740, 129–137 (1994).

11. Chen-Germán, M. et al. Detection of Oropouche and Punta Toro Virus Infections by Enhanced Surveillance, Panama, 2023–2024. Emerg. Infect. Dis. 32, (2026).

12. Naveca, F. G. et al. Human outbreaks of a novel reassortant Oropouche virus in the Brazilian Amazon region. Nat. Med. 30, 3509–3521 (2024).

13. Scachetti, G. C. et al. Re-emergence of Oropouche virus between 2023 and 2024 in Brazil: an observational epidemiological study. Lancet Infect. Dis. 25, 166–175 (2025).

14. Manuli, E. R. et al. Transmission dynamics of Oropouche virus in Latin America and the Caribbean. Nat. Med. 32, 1383–1392 (2026).

15. Hua, X. et al. Ecological and demographic drivers of Oropouche virus transmission. *Nat*. Health 1, 487–496 (2026).

16. Tegally, H. et al. Dynamics and ecology of a multistage expansion of Oropouche virus in Brazil. Nat. Ecol. Evol. https://doi.org/10.1038/s41559-026-03042-0 (2026) doi:10.1038/s41559-026-03042-0.

17. Pan American Health Organization. Increased migration flow in the Americas in 2023: Challenges for migrant health and PAHO’s response. https://www.paho.org/en/news/18-12-2023-increased-migration-flow-americas-2023-challenges-migrant-health-and-pahos-response (2023).

18. Oidtman, R. J., España, G. & Perkins, T. A. Co-circulation and misdiagnosis led to underestimation of the 2015–2017 Zika epidemic in the Americas. PLoS Negl. Trop. Dis. 15, e0009208 (2021).

19. Tilston-Lunel, N. L. et al. Genetic analysis of members of the species Oropouche virus and identification of a novel M segment sequence. J. Gen. Virol. 96, 1636–1650 (2015).

20. Gutierrez, B. et al. Evolutionary Dynamics of Oropouche Virus in South America. J. Virol. 94, e01127–19 (2020).

21. Mercer, D. R., Spinelli, G. R., Watts, D. M. & Tesh, R. B. Biting Rates and Developmental Substrates for Biting Midges (Diptera: Ceratopogonidae) in Iquitos, Peru. J. Med. Entomol. 40, 807–812 (2003).

22. Feitoza, L. H. M. et al. Influence of meteorological and seasonal parameters on the activity of Culicoides paraensis (Diptera: Ceratopogonidae), an annoying anthropophilic biting midge and putative vector of Oropouche Virus in Rondônia, Brazilian Amazon. Acta Trop. 243, 106928 (2023).

23. Gräf, T. et al. Expansion of Oropouche virus in non-endemic Brazilian regions: analysis of genomic characterisation and ecological drivers. Lancet Infect. Dis. 25, 379–389 (2025).

24. Shahid, F. et al. The emerging threat of Oropouche virus in Latin America: epidemiology, clinical manifestations, diagnosis, and management. Ann. Med. Surg. 87, 8391–8399 (2025).

25. Milani, P. et al. Molecular and Serological Evidence of Oropouche Virus Circulation in Asymptomatic Blood Donors During the 2023–2024 Outbreak in Manaus, Brazil. J. Infect. Dis. jiag088 (2026) doi:10.1093/infdis/jiag088.

26. Carson Institute. The Darién Gap Cohort. https://carsoninstitute.org/2024/10/24/the-darien-gap-cohort/.

27. FilmArray® Global Fever RUO Panel. Biofire Defense LLC – B2B store 1 https://biofiredefense.com/filmarray-global-fever-ruo-panel/.

28. Pan American Health Organization. Oropouche virus disease. https://www.paho.org/en/topics/oropouche-virus-disease (2025).

29. Rojas, A. et al. Real-time RT-PCR for the detection and quantitation of Oropouche virus. Diagn. Microbiol. Infect. Dis. 96, 114894 (2020).

30. Sabalza, M. et al. A-282 Detection of anti-Oropouche virus-specific IgM in a patient panel from a Zika virus epidemic area in 2015 / 2016. Clin. Chem. 71, hvaf086.271 (2025).

31. Ma, J. & Earn, D. J. D. Generality of the Final Size Formula for an Epidemic of a Newly Invading Infectious Disease. Bull. Math. Biol. 68, 679–702 (2006).

32. Murrell, B. et al. Detecting Individual Sites Subject to Episodic Diversifying Selection. PLoS Genet. 8, e1002764 (2012).

33. Murrell, B. et al. FUBAR: A Fast, Unconstrained Bayesian AppRoximation for Inferring Selection. Mol. Biol. Evol. 30, 1196–1205 (2013).

34. Smith, M. D. et al. Less Is More: An Adaptive Branch-Site Random Effects Model for Efficient Detection of Episodic Diversifying Selection. Mol. Biol. Evol. 32, 1342–1353 (2015).

35. Korber, B. & Myers, G. Signature Pattern Analysis: A Method for Assessing Viral Sequence Relatedness. AIDS Res. Hum. Retroviruses 8, 1549–1560 (1992).

