## Supplementary Materials for "Integrated surveillance resolves Darién paradox of Oropouche virus emergence in Panama’s migration corridor"

**Methods**

**Statistical analysis for clinical symptoms and risk factors**

Cytokine analyses were performed using the LEGENDplex panel on 97 serum samples obtained from permanent residents of Darién Province, Panama, including 56 RT-qPCR-confirmed OROV-positive individuals and 41 OROV-negative healthy controls. Controls were frequency-matched by age range and sex. OROV-positive participants were recruited from healthcare centers between December 2024 and March 2025 and presented with mild disease and ≤7 days since symptom onset, whereas healthy controls corresponded to samples collected during a cross-sectional study conducted in Darién in 2018. No samples were excluded due to insufficient volume or poor sample quality. Rather, the final sample size analyzed was determined by the limited availability and high cost of the multiplex cytokine panel reagents. Subsequent bivariate regression analyses were adjusted for age and sex.

To estimate adjusted Relative Risk (aRRs), generalized linear models (GLMs) with a Poisson distribution, log link function, and robust variance estimation were applied. Statistical significance was defined as a p-value < 0.05, and 95% confidence intervals (95% CI) were reported for all estimates. This sub-analysis included a subset of participants (n=289) rather than the full study population, identified through household visits in communities with the highest case burden during the outbreak; enrolment was voluntary following informed explanation, with eligibility restricted to individuals aged 4–70 years in accordance with the approved bioethics protocol.

To compare clinical presentations between OROV infection and other endemic arboviruses, separate binary outcomes were defined for each comparison. In each model, OROV infection was coded as 1, and the comparator virus was coded as 0 (DENV, ZIKV, CHIKV, or MADV/VEEV). Variable selection was conducted using a nested model approach based on likelihood ratio tests and p-value thresholds. Multivariable analyses were performed using generalized linear models with a Poisson distribution, log link, and robust variance estimation to estimate adjusted Relative Risk (aRRs). Model building followed a forward stepwise approach with nested models, using log-likelihood comparisons for variable selection. Variables with two-sided p values <0.05 were retained in the final models. Effect estimates are presented with 95% confidence intervals.

**Within household transmission method**

We reconstructed household membership using geospatial coordinates recorded for each participant. A deterministic household identifier (house_code) was created by hashing the concatenated latitude and longitude strings, ensuring that all individuals sharing identical coordinates were assigned to the same dwelling. Household size was computed as the total number of unique individuals linked to each identifier. Households were eligible for analysis if they contained ≥2 members and ≥1 individual classified as infected.

To capture both symptomatic and subclinical OROV infections, infection status was defined using a composite laboratory-based definition. Individuals were classified as infected if they tested positive by PCR, IgM ELISA, IgG ELISA, ELISA among household contacts, or PRNT.

Within-household transmission was quantified using a chain binomial final-size model^2^. The model assumes that a single infection is introduced into a household, each susceptible individual becomes infected with a constant probability $p$, independent of other individuals, and that transmission proceeds until no susceptible members remain.

For each household $h$:

$n_{h}$: number of household members

$z_{h}$: number of individuals classified as infected

Assuming a single primary introduction per household, the number of secondary infections was defined as:

$$I_{2,h}=max(z_{h}-1,0),$$

and the number of individuals initially susceptible was:

$$S_{h}=n_{h}-1.$$

The pooled estimate of the household secondary attack probability was calculated as:

$$\hat{p}_{final}=\frac{\sum_{h} I_{2,h}}{\sum_{h} S_{h}}.$$

Exact 95% confidence intervals (CI) were computed using the Clopper–Pearson method for binomial proportions. Because this approach relies on final infection status rather than temporal ordering, it is well suited for pathogens with a high proportion of asymptomatic or retrospectively identified infections, including orthobunyaviruses and other arboviruses^3^.

**Risk Mapping**

Municipal/Corregimiento Data Acquisition and Processing

The fitting of the risk model was performed at the Brazilian municipal level. Brazilian municipal populations were provided by the 2022 Brazilian population censuses reported by the Brazilian Institute of Geography and Statistics. See Table S10 for sources of features including human density, typical climate, land use for specific crops, human footprint, and soil conditions, and time-varying weather conditions.

Shapefiles for Panamanian corregimientos were obtained using the `geodata`R package^4^. All data in raster format were extracted to the municipal/corregimiento level using the R package ‘exactextractr’ version 0.10.0^5^   and 2020 municipality boundaries provided by  the R package ‘geobr’ version 1.9.0^6^. To reflect the population density experienced by humans, we used a weighted mean of population density, weighted by population density. For other features, an unweighted mean was obtained. Risk models were fit using the mean for each weather condition in the 30 days before and after each introduction. All predictors were z-score normalized prior to fitting.

Outbreak Determination

For each municipality, sample collection dates for OROV-positive individuals were converted into daily incidence using the R package ‘incidence’ version 1.7.5^7^. Cases were grouped into periods, with cases separated by 21 days or fewer (by sample collection date) considered to be in the same period. Periods with fewer than three cases were considered isolated cases, and periods with three or more cases were considered outbreaks. Introduction date is the date of an isolated case, or the date of the first case in an outbreak.

Statistical analysis

We used a generalized linear modeling (GLM) framework to identify features and weather conditions of municipalities where introduction resulted in (i) outbreak or (ii) no outbreak.

Variable selection was performed in two stages. For initial variable selection, we applied a random forest approach using R package ‘randomForestSRC’ version 3.3.3^8^ for variable ranking and selection of factors to include in the next step of the regression analysis, which were used to fit a GLM. That model was further reduced using stepwise regression (R package ‘stats’ version 4.4.2^9^). Finally, the model was further reduced by manual removal of predictors, where removal caused a reduction or small (<2) increase in AIC. We arrived at the Minimum Adequate Model (MAM) by removing in order variables as listed in Table S12. The AIC of the MAM is 1060.3.  See Table S11 for all model AIC and Δ AIC values. See Figure S13	for visualization of variable importance and Figure S14	for the Odds Ratio (OR) associated with each variable.

Map Generation

To produce risk projections within Panama, all predictors were normalized using the mean and standard deviations of each predictor within Brazil (as these values were used to z score normalize the model fit within Brazil). Five corregimientos, all small islands, did not have soil data available; all were assigned the mean value for each predictor. “Typical” risk for each month was determined using time-invariant features and monthly climate normals in lieu of weather. To produce Figure 7A, Figure 7B, and Supplemental Figure S19 Climate Panels, weather for each month was derived from climate normals. The monthly climate normals used are expected average values for each month of the daily average vapor pressure, daily minimum, maximum, and mean temperatures, and total precipitation, based on historic (1970-2000) weather data^10^. To produce Figure 7C and Supplemental Figure 19 panels 2023-2025 risk was calculated using weather conditions observed in each month of 2023, 2024, and 2025.

**RNA extraction and sequencing**

Total RNA was extracted from 200 µL of samples using 600 µL of TRIzol™ LS Reagent (Thermo Fisher Scientific, 10296010), following the manufacturer’s instructions. Shotgun short-read libraries were prepared using the NEBNext® Ultra™ II RNA Library Prep Kit for Illumina (New England Biolabs, Ipswich, MA, USA). Briefly, 26 µL of RNA (2–4 ng/µL) was enzymatically fragmented, followed by ligation of hairpin sequencing adapters containing 5′-dT overhangs and a uridine ribonucleotide within the hairpin loop, which was subsequently cleaved at the uridine site. Libraries were PCR-amplified and indexed using NEBNext® Unique Dual Indexes.

Library quality was assessed using the Agilent D1000 ScreenTape assay (Agilent Technologies, Santa Clara, CA, USA). Libraries passing quality control were quantified using the Qubit™ dsDNA BR Assay Kit (Thermo Fisher Scientific), pooled, and sequenced on an Illumina NovaSeq 6000 platform using S4 and SP flow cells (Illumina, San Diego, CA, USA).

**Bioinformatics and data cleaning**

Raw sequencing reads generated on the NovaSeq 6000 were quality-filtered and trimmed using fastp v0.23.4, applying the following criteria: Phred score ≥30, ≥40% of bases above Phred 30, average paired-read quality ≥30, and minimum read length ≥50 bp. Trimmed reads were processed using the BDRD in-house *All-in-One* pipeline for taxonomic classification and de novo assembly, with assemblies generated using metaSPAdes v4.0.0.

Both trimmed reads and assembled contigs were aligned for sequence identification using MMSeqs2 v15.6f452 for nucleotide-level comparisons against the NCBI *core_nt* database and DIAMOND v2.1.9 for protein-level comparisons against the NCBI *nr* database. Resulting DIAMOND alignment files (.daa) were imported into MEGAN v7.1.1 for taxonomic visualization and assessment.

Read mapping was performed to validate organism detection and estimate genomic coverage using BBTools v36.06. Reads were aligned with BBMap, setting the maximum insertion/deletion length to 50 bp, and ambiguous reads were resolved using BBSplit by assigning them to the best-matching reference. De novo assembled OROV contigs were extracted and imported into CLC Genomics Workbench v25.0.3 for downstream analyses. Open reading frames were identified and translated, followed by pairwise sequence comparisons to assess percent identity and amino acid differences.

**Molecular signature, recombination, phylogenetics and phylogeographic analysis**Full genomes (n=613 for L segment, n=691 for M segment, and n=647 for S segment) with geographical location, time of isolation, and source were downloaded from GenBank (Table S4). Alignment of the full-genomes sequences, segments of the genome (S, M, and L) corresponding to recombinant regions were visualized, inspected and removed using RPD5 with six methods BOOTSCAN/RECSCAN, GENECONV, MAXCHI, CHIMAERA, 3SEQ, SISCAN. To identify sites and branches under positive selection, we used the hyphy package implemented through the Datamonkey web server (https://www.datamonkey.org/) to perform three complementary analyses: 1. Mixed effects model of evolution^11^ (MEME) detects diversifying selection at individual sites, allowing the distribution of ω (dN/dS) to vary across sites and branches. 2. Fast Unconstrained Bayesian AppRoximation^12^ (FUBAR) to identify sites subject to pervasive diversifying or purifying selection using a Bayesian approach. 3. Adaptive Branch-Site Random Effects Likelihood^13^ (aBSREL) to test for episodic diversifying selection on branches, allowing ω to vary across branches and sites. Sites were considered to have experienced statistically significant positive or negative selection based on the following cut-offs: LRT p ≤ 0.05 for aBSREL and posterior probability (PP) > 0.90 for FUBAR, and LRT ≤ 0.05 for MEME. Viral epidemiology signature pattern analysis was used to identify and estimate the frequency of each amino acid at each position across the OROV viral alignment, using Panama isolates as query and regional isolates as background^14^. Ancestral sequence reconstruction was performed using Treetime v0.11.4 to identify branch mutations along the tree for each OROV segment. Maximum likelihood was carried out to determine the phylogenetic signal for each segment using IQTree v2.4.0. To evaluate temporal signal in each viral segment alignment, root-to-tip regression analysis was conducted using TempEst v1.5.3^15^ and Treetime v0.11.4^16^, using a heuristic residual mean squared optimization approach to assess the correlaction between genetic divergence (root to tip distance) and sampling time, therefore determining whether sufficient molecular clock signal is present for reliable phylogenetic dating^15^. This analysis assessed the correlation between sampling dates and genetic divergence from the root, with statistical significance determined by Pearson correlation coefficient to determine suitability for molecular clock calibration. Ancestral sequence reconstruction was performed using Treetime v0.11.4 to identify branch mutations along the tree for each OROV segment. The spatiotemporal spread of OROV strains from 2020 to 2025 was analyzed using a phylogeographic reconstruction with diffusion in discrete space implemented in BEAST v10.5.0. Bayesian trees were reconstructed using GTR+G4+I nucleotide substitution model, uncorrelated relaxed clock, and Hamiltonian Monte Carlo SkyGrid. An asymmetric model with a Bayesian stochastic search variable selection (BSSVS) was implemented to identify the important migration routes using a discrete location that were assigned according to the sampling township only for Panama (PAN:Cemaco, PAN:El_real, PAN:Lajas_Blanca, PAN:Meteti, PAN:Rio_Iglesia, PAN:Sambu, PAN:Yape, PAN:Yaviza). We performed multiple 250 million MCMC runs to ensure ESS >100-200, which were visualized in Tracer v1.7. The maximum clade credibility (MCC) trees were summarized with TreeAnnotator v10.5.0 and visualized using FigTree v.1.4.5. Bayes factors (BF) to summarize migratory events and to identify statistically supported migration routes (BF > 5) were calculated in R/Rstudio packages. ML and Bayesian time-based trees were plotted using R/Rstudio packages.

**Supplementary Tables and Figures**

Tables

**Table S1.** Symptom profile and statistical comparison between OROV-negative and OROV-positive participants.

| Symptom | Category | N | % | Negative n=613 | Positive n=447 | p^a^ | RR | RR  95% CI | p^b^ |
| --- | --- | --- | --- | --- | --- | --- | --- | --- | --- |
| Fever | |  |  |  |  | <0.001 |  |  |  |
|  | No | 254 | 24.0 | 117 (47.4) | 130 (52.6) |  | Ref. |  |  |
|  | Yes | 806 | 76.0 | 476 (60.0) | 317 (40.0) |  | 0.77 | 0.68 - 0.89 | 0.001 |
| Headache | |  |  |  |  | 0.789 |  |  |  |
|  | No | 304 | 28.7 | 169 (56.3) | 131 (43.7) |  | Ref. |  |  |
|  | Yes | 755 | 71.3 | 423 (57.2) | 316 (42.8) |  | 0.98 | 0.84 - 1.15 | 0.835 |
| Chills | |  |  |  |  | 0.186 |  |  |  |
|  | No | 677 | 63.9 | 391 (58.50 | 277 (41.5) |  | Ref. |  |  |
|  | Yes | 383 | 36.1 | 202 (54.3) | 170 (45.7) |  | 1.09 | 0.95 - 1.26 | 0.223 |
| Myalgia | |  |  |  |  | 0.005 |  |  |  |
|  | No | 654 | 61.8 | 388 (60.3) | 255 (39.7) |  | Ref. |  |  |
|  | Yes | 405 | 38.2 | 204 (51.5) | 192 (48.5) |  | 1.21 | 1.05 - 1.39 | 0.009 |
| Arthralgia | |  |  |  |  | 0.088 |  |  |  |
|  | No | 702 | 66.2 | 408 (58.9) | 285 (41.1) |  | Ref. |  |  |
|  | Yes | 358 | 33.8 | 185 (53.3) | 162 (46.7) |  | 1.12 | 0.96 - 1.28 | 0.138 |
| Retroorbital pain | |  |  |  |  | 0.053 |  |  |  |
|  | No | 830 | 78.3 | 478 (58.6) | 338 (41.4) |  | Ref. |  |  |
|  | Yes | 230 | 21.7 | 115 (51.3) | 109 (48.7) |  | 1.17 | 1.00 - 1.37 | 0.049 |
| Diarrhea | |  |  |  |  | 0.708 |  |  |  |
|  | No | 967 | 91.2 | 540 (56.8) | 410 (43.2) |  | Ref. |  |  |
|  | Yes | 93 | 8.8 | 53 (58.9) | 37 (41.1) |  | 0.95 | 0.73 - 1.22 | 0.671 |
| Rash | |  |  |  |  | <0.001 |  |  |  |
|  | No | 1020 | 96.4 | 582 (58.1) | 420 (41.9) |  | Ref. |  |  |
|  | Yes | 38 | 3.6 | 9 (25.0) | 27 (75.0) |  | 1.77 | 1.44 - 2.16 | <0.001 |
| Conjunctivitis | |  |  |  |  | 0.421 |  |  |  |
|  | No | 805 | 99.3 | 522 (65.7) | 273 (34.3) |  | Ref. |  |  |
|  | Yes | 6 | 0.7 | 3 (50.0) | 3 (50.0) |  | 1.40 | 0.59 - 3.34 | 0.446 |
| Abdominal pain | |  |  |  |  | 0.006 |  |  |  |
|  | No | 996 | 94.0 | 569 (58.1) | 410 (41.9) |  | Ref |  |  |
|  | Yes | 63 | 6.0 | 24 (40.0) | 36 (60.0) |  | 1.41 | 1.14 - 1.75 | 0.002 |
| Lumbar pain | |  |  |  |  | 0.392 |  |  |  |
|  | No | 785 | 96.8 | 510 (65.8) | 265 (34.2) |  | Ref. |  |  |
|  | Yes | 26 | 3.2 | 15 (57.7) | 11 (42.3) |  | 1.20 | 0.75 - 1.91 | 0.445 |
| Nausea or Vomiting | |  |  |  |  | 0.253 |  |  |  |
|  | No | 740 | 69.8 | 405 (55.9) | 320 (44.1) |  | Ref. |  |  |
|  | Yes | 320 | 30.2 | 188 (59.7) | 127 (40.3) |  | 0.91 | 0.78 - 1.06 | 0.231 |
| Hepatomegaly | |  |  |  |  | 0.643 |  |  |  |
|  | No | 809 | 99.8 | 524 (65.6) | 275 (34.4) |  | Ref. |  |  |
|  | Yes | 2 | 0.2 | 1 (50.0) | 1 (50.0) |  | 1.52 | 0.37 - 6.28 | 0.562 |
| Hemorrhage | |  |  |  |  | 0.737 |  |  |  |
|  | No | 1056 | 99.7 | 591 (57.0) | 445 (43.0) |  | Ref. |  |  |
|  | Yes | 3 | 0.3 | 2 (66.7) | 1 (33.3) |  | 0.78 | 0.16 - 3.92 | 0.769 |
| Appetite loss | |  |  |  |  | 0.548* |  |  |  |
|  | No | 809 | 99.8 | 523 (65.5) | 276 (34.5) |  |  |  |  |
|  | Yes | 2 | 0.2 | 2 (100.0) | 0(0.0) |  |  |  |  |
| Seizure | |  |  |  |  | 0.775 |  |  |  |
|  | No | 1055 | 99.6 | 591 (57.1) | 444 (42.9) |  | Ref. |  |  |
|  | Yes | 2 | 0.4 | 2(50.0) | 2(50.0) |  | 1.09 | 0.40 - 2.93 | 0.865 |

N, total number of participants

%, percentage

RR, Relative Risk.

95% CI, 95% Confidence Interval.

^a^Chi-square test p value.

^b^Bivariable Poisson regression (p value); adjusted for sex and age.

* Fisher’s exact test p value.

**Table S2.** Overview of the study population and bivariate assessment of factors associated with infection (n=289).

| Variables | Categories |  | N | % | Negative (n=67) | Positive (n=222) | *95% CI | ^a^p-value |
| --- | --- | --- | --- | --- | --- | --- | --- | --- |
| Age (Quintile) | |  |  |  |  |  |  | 0.025 |
|  | Q1 (0-16 years) | | 62 | 21.5 | 20 (32.3) | 42 (67.7) | 54.7 - 79.1 |  |
|  | Q2 (17-27 years) | | 58 | 20.1 | 10 (17.2) | 48 (82.8) | 70.6 - 91.4 |  |
|  | Q3 (28-37 years) | | 57 | 19.8 | 13 (22.8) | 44 (77.2) | 64.2 - 87.3 |  |
|  | Q4 (38-53 years) | | 55 | 19.1 | 6 (10.9) | 49 (89.1) | 77.8 - 95.9 |  |
|  | Q5 (54-75 years) | | 56 | 19.4 | 18 (32.1) | 38 (67.9) | 54.0 - 79.7 |  |
| Sex |  |  |  |  |  |  |  | 0.134 |
|  | Male |  | 107 | 37.0 | 30 (28.0) | 77 (72.0) | 62.4 - 80.2 |  |
|  | Female |  | 182 | 63.0 | 37 (20.3) | 145 (79.7) | 73.1 - 85.3 |  |
| Length of residence (Tercil) | |  |  |  |  |  |  | 0.384 |
|  | T1 (≤4 years) | | 98 | 34.0 | 25 (25.1) | 73 (74.5) | 64.7 - 82.8 |  |
|  | T2 (5-19 years | | 97 | 33.7 | 25 (25.7) | 72 (74.2) | 64.4 - 82.6 |  |
|  | T3 (≥20 years) | | 93 | 32.3 | 17 (18.3) | 76 (81.7) | 72.4 - 89.0 |  |
| Ethnicity |  |  |  |  |  |  |  | 0.230 |
|  | Mestizo |  | 153 | 53.3 | 33 (21.6) | 120 (78.4) | 71.1 - 84.7 |  |
|  | Indigenous |  | 50 | 17.4 | 11 (22.0) | 39 (78.0) | 64.0 - 88.5 |  |
|  | Afro-descendant | | 29 | 10.1 | 10 (34.5) | 19 (65.5) | 45.7 - 82.1 |  |
|  | Caucasian |  | 52 | 18.1 | 11 (21.2) | 41 (78.8) | 65.3 - 88.9 |  |
|  | Other |  | 3 | 1.0 | 2 (66.7) | 1 (33.3) | 0.8 - 90.6 |  |
| Education level | |  |  |  |  |  |  | 0.688 |
|  | None |  | 21 | 7.4 | 7 (33.3) | 14 (66.7) | 43.0 - 85.4 |  |
|  | Primary |  | 112 | 39.2 | 24 (21.4) | 88 (78.6) | 69.8 - 85.8 |  |
|  | Secondary |  | 109 | 38.1 | 25 (22.9) | 84 (77.1) | 68.0 - 84.6 |  |
|  | Technical/University | | 44 | 15.4 | 11 (25.0) | 33 (75.0) | 59.7 - 86.8 |  |
| Family income (dollar) | |  |  |  |  |  |  | 0.744 |
|  | ≤ 400 |  | 187 | 67.0 | 48 (25.7) | 139 (74.3) | 67.4 - 80.4 |  |
|  | >400 & <700 | | 48 | 17.2 | 11 (22.9) | 37 (77.1) | 62.7 - 88.0 |  |
|  | ≥700 & <1000 | | 19 | 6.8 | 3 (13.8) | 16 (84.2) | 60.4 - 96.6 |  |
|  | ≥1000 |  | 25 | 9.0 | 5 (20.0) | 20 (80.0) | 59.3 - 93.2 |  |
| Floor type |  |  |  |  |  |  |  | 0.310 |
|  | Dirt |  | 27 | 9.4 | 7 (25.9) | 20 (74.1) | 53.7 - 88.9 |  |
|  | Wooden |  | 17 | 5.9 | 2 (11.8) | 15 (88.2) | 63.6 - 98.5 |  |
|  | Tiled |  | 63 | 21.9 | 19 (30.2) | 44 (69.8) | 57.0 - 80.8 |  |
|  | Concrete |  | 181 | 62.8 | 38 (21.0) | 143 (79.0) | 72.3 - 84.7 |  |
| Wall type |  |  |  |  |  |  |  | 0.433 |
|  | Concrete |  | 170 | 59.4 | 38 (22.4) | 132 (77.6) | 70.6 - 83.7 |  |
|  | Wooden |  | 106 | 37.1 | 24 (22.6) | 82 (77.4) | 68.2 - 84.9 |  |
|  | Other |  | 10 | 3.5 | 4 (40.0) | 6 (60.0) | 26.2 - 87.8 |  |
| Room quantity | |  |  |  |  |  |  | 0.061 |
|  | ≤2 |  | 161 | 55.7 | 44 (27.3) | 117 (72.7) | 65.1 - 79.4 |  |
|  | >2 |  | 128 | 44.3 | 23 (18.0) | 105 (82.0) | 74.3 - 88.2 |  |
| Bathroom type | |  |  |  |  |  |  | 0.009 |
|  | Flush toilet |  | 194 | 68.3 | 53 (27.3) | 141 (72.7) | 65.8 - 78.8 |  |
|  | Latrine |  | 90 | 31.7 | 12 (13.3) | 78 (86.7) | 77.9 - 92.9 |  |
| Garbage disposal | |  |  |  |  |  |  | 0.249 |
|  | Garbage truck | | 144 | 51.1 | 38 (26.39) | 106 (73.6) | 65.6 - 80.6 |  |
|  | Burned |  | 110 | 39.0 | 20 (18.2) | 90 (81.8) | 73.3 - 88.5 |  |
|  | Buried / Other | | 28 | 9.9 | 5 (17.9) | 23 (82.1) | 63.1 - 93.9 |  |
| Water source | |  |  |  |  |  |  | 0.937 |
|  | Public aqueduct | | 249 | 88.6 | 56 (22.5) | 193 (77.5) | 71.8 - 82.5 |  |
|  | Other sources | | 32 | 11.4 | 7 (21.9) | 25 (78.1) | 60.0 - 90.7 |  |
| Water storage | |  |  |  |  |  |  | 0.581 |
|  | No |  | 71 | 25.3 | 14 (19.7) | 57 (80.3) | 69.1 - 88.8 |  |
|  | Yes |  | 210 | 74.7 | 48 (22.9) | 162 (77.1) | 70.9 - 82.6 |  |
| Type of water receptacle | |  |  |  |  |  |  | 0.775 |
|  | Plastic |  | 198 | 88.0 | 49 (24.8) | 149 (75.2) | 68.6 - 81.1 |  |
|  | Reserve tank / Other | | 27 | 12.0 | 6 (22.2) | 21 (77.8) | 57.7 - 91.4 |  |
| Does the water receptacle have a lid | | |  |  |  |  |  | 0.244 |
|  | No |  | 42 | 18.3 | 13 (31.0) | 29 (69.0) | 52.9 - 82.4 |  |
|  | Yes |  | 187 | 81.7 | 42 (22.5) | 145 (77.5) | 70.9 - 83.3 |  |
| Main Occupation | |  |  |  |  |  |  | 0.820 |
|  | Housewife |  | 110 | 40.3 | 22 (20.0) | 88 (80.0) | 71.3 - 87.0 |  |
|  | Farmer / cattle rancher | | 19 | 7.0 | 5 (26.3) | 14 (73.7) | 48.8 - 90.8 |  |
|  | Student |  | 75 | 27.5 | 19 (25.3) | 56 (74.7) | 63.3 - 84.0 |  |
|  | Other |  | 69 | 25.3 | 15 (21.7) | 54 (78.3) | 66.7 - 87.3 |  |

N, total number of participants

%, percentage

*95% CI, 95% confidence interval for incident cases.

^a^Chi-square test p value.

**Table S3.** Poisson regression analysis of infection risk factors: bivariate and multivariable models adjusted for sex and age.

| Variables | Categories | RR | 95% CI | ^a^p-value | RR | 95% CI | ^b^p-value |
| --- | --- | --- | --- | --- | --- | --- | --- |
| Age (Continue) | |  |  |  | 1.00 | 0.99 - 1.01 | 0.819 |
| Age (Quintile) | |  |  |  |  |  |  |
|  | Q1 (0-16 years) | Ref. |  |  |  |  |  |
|  | Q2 (17-27 years) | 1.22 | 0.99 - 1.50 | 0.060 |  |  |  |
|  | Q3 (28-37 years) | 1.14 | 0.91 - 1.42 | 0.250 |  |  |  |
|  | Q4 (38-53 years) | 1.32 | 1.08 - 1.60 | 0.006 |  |  |  |
|  | Q5 (54-75 years) | 1.00 | 0.78 - 1.28 | 0.989 |  |  |  |
| Sex |  |  |  |  |  |  |  |
|  | Male | Ref. |  |  | Ref. |  |  |
|  | Female | 1.11 | 0.96 - 1.27 | 0.153 | 1.19 | 0.94 - 1.52 | 0.147 |
| Length of residence (Tercil) | |  |  |  |  |  |  |
|  | Q1 (≤4 years) | Ref. |  |  |  |  |  |
|  | Q1 (5-19 years | 0.98 | 0.82 - 1.51 | 0.783 |  |  |  |
|  | Q1 (≥20 years) | 1.12 | 0.94 - 1.32 | 0.205 |  |  |  |
| Ethnicity |  |  |  |  |  |  |  |
|  | Mestizo | Ref. |  |  | Ref. |  |  |
|  | Indigenous | 0.98 | 0.83 - 1.17 | 0.861 | 0.89 | 0.71 - 1.12 | 0.318 |
|  | Afro-descendant | 0.84 | 0.64 - 1.11 | 0.215 | 0.78 | 0.56 - 1.08 | 0.131 |
|  | Caucasian | 1.00 | 0.85 - 1.18 | 0.957 | 1.02 | 0.84 - 1.23 | 0.849 |
|  | Other | 0.43 | 0.08 - 2.21 | 0.316 |  |  |  |
| Education level | |  |  |  |  |  |  |
|  | None | 0.88 | 0.62 - 1.26 | 0.481 |  |  |  |
|  | Primary | 1.05 | 0.86 - 1.28 | 0.624 |  |  |  |
|  | Secondary | 1.03 | 0.84 - 1.26 | 0.797 |  |  |  |
|  | Technical/University | Ref. |  |  |  |  |  |
| Family income | |  |  |  |  |  |  |
|  | ≤ 400 | Ref. |  |  |  |  |  |
|  | >400 & <700 | 1.04 | 0.87 - 1.25 | 0.628 |  |  |  |
|  | ≥700 & <1000 | 1.17 | 0.94 - 1.45 | 0.149 |  |  |  |
|  | ≥1000 | 1.11 | 0.88 - 1.40 | 0.363 |  |  |  |
| Floor type |  |  |  |  |  |  |  |
|  | Dirt | Ref. |  |  |  |  |  |
|  | Wooden | 1.19 | 0.89 - 1.58 | 0.241 |  |  |  |
|  | Tiled | 0.93 | 0.70 - 1.24 | 0.627 |  |  |  |
|  | Concrete | 1.06 | 0.83 - 1.43 | 0,656 |  |  |  |
| Wall type |  |  |  |  |  |  |  |
|  | Concrete | Ref. |  |  |  |  |  |
|  | Wooden | 1.0 | 0.87 - 1.13 | 0.977 |  |  |  |
|  | Other | 0.78 | 0.46 - 1.33 | 0.364 |  |  |  |
| Room quantity | |  |  |  |  |  |  |
|  | ≤2 | Ref. |  |  |  |  |  |
|  | >2 | 1.13 | 0.99 - 1.28 | 0.062 |  |  |  |
| Bathroom type | |  |  |  |  |  |  |
|  | Flush toilet | Ref. |  |  | Ref. |  |  |
|  | Latrine | 1.20 | 1.07 - 1.35 | 0.003 | 1.16 | 0.99 - 1.35 | 0.06 |
| Garbage disposal | |  |  |  |  |  |  |
|  | Garbage truck | Ref. |  |  | Ref. |  |  |
|  | Burned | 1.10 | 0.96 - 1.26 | 0.147 | 1.18 | 1.01 - 1.39 | 0.037 |
|  | Buried / Other | 1.10 | 0.90 - 1.36 | 0.350 | 1.17 | 0.92 - 1.48 | 0.200 |
| Water source | |  |  |  |  |  |  |
|  | Public aqueduct | Ref. |  |  | Ref. |  |  |
|  | Other sources | 1.01 | 0.82 - 1.23 | 0.936 | 1.05 | 0.86 - 1.28 | 0.652 |
| Water storage | |  |  |  |  |  |  |
|  | No | Ref. |  |  |  |  |  |
|  | Yes | 0.96 | 0.84 - 1.10 | 0.589 |  |  |  |
| Type of water receptacle | |  |  |  |  |  |  |
|  | Plastic | Ref. |  |  | Ref. |  |  |
|  | Reserve tank / Other | 1.02 | 0.82 - 1.27 | 0.857 | 1.09 | 0.86 - 1.37 | 0.479 |
| Does the water receptacle have a lid | |  |  |  |  |  |  |
|  | No | Ref. |  |  |  |  |  |
|  | Yes | 1.14 | 0.91 - 1.42 | 0.247 |  |  |  |
| Main Occupation | |  |  |  |  |  |  |
|  | Housewife | Ref. |  |  | Ref. |  |  |
|  | Farmer / cattle rancher | 1.01 | 0.72 - 1.42 | 0.947 | 1.01 | 0.65 - 1.57 | 0.952 |
|  | Student | 0.88 | 0.72 - 1.09 | 0.246 | 0.98 | 0.77 - 1.26 | 0.905 |
|  | Other | 1.01 | 0.80 - 1.26 | 0.952 | 1.28 | 0.99 - 1.64 | 0.055 |

^a^Bivariable Poisson regression (p value); adjusted for sex and age.

^b^Multivariable Poisson regression (p value); adjusted for sex and age.

RR, Relative risk

95% CI, 95% confidence interval

**Table S4.** Characteristics of cases and contacts and bivariate analysis of associated factors.

| Variables | Categories | N | % | Case (n=47) | Contact (n=242) | ^a^p-value |
| --- | --- | --- | --- | --- | --- | --- |
| Knows sick person outside home | |  |  |  |  | 0.992 |
|  | No | 61 | 21.3 | 10 (16.4) | 51 (83.6) |  |
|  | Yes | 225 | 78.7 | 37 (16.4) | 188 (83.6) |  |
| Sick Contact | |  |  |  |  | 0.607 |
|  | No | 61 | 21.6 | 11 (18.0) | 50 (82.0) |  |
|  | Yes | 222 | 78.4 | 34 (15.4) | 188 (84.7) |  |
| Sick contact location | |  |  |  |  | 0.018 |
|  | At household | 68 | 30.8 | 10 (14.7.0) | 58 (85.3) |  |
|  | Outside household | 81 | 36.6 | 19 (23.5) | 62 (76.5) |  |
|  | Both | 72 | 32.6 | 5 (6.94) | 67 (93.1) |  |
| Sick contact date | |  |  |  |  | 0.082 |
|  | February 2025 | 99 | 44.6 | 20 (20.2) | 79 (79.8) |  |
|  | January 2025 | 89 | 40.1 | 8 (9.0) | 81 (91.0) |  |
|  | Sep-Dec 2024 | 34 | 15.3 | 4 (11.8) | 30 (88.2) |  |
| Internal movement | |  |  |  |  | 0.446 |
|  | No | 155 | 54.0 | 23 (14.8) | 132 (85.2) |  |
|  | Yes | 132 | 46.0 | 24 (18.2) | 108 (81.8) |  |
| External movement | |  |  |  |  | 0.080 |
|  | No | 188 | 65.5 | 36 (19.2) | 152 (80.8) |  |
|  | Yes | 99 | 34.5 | 11 (11.1) | 88 (88.9) |  |
| Fever in the last month | |  |  |  |  | 0.003 |
|  | No | 90 | 31.2 | 6 (6.7) | 84 (93.3) |  |
|  | Yes | 198 | 68.8 | 41 (20.7) | 157 (79.3) |  |
| Fever Date |  |  |  |  |  | 0.001 |
|  | Feb-2025 | 87 | 43.9 | 29 (33.3) | 58 (66.7) |  |
|  | Jan-2025 | 89 | 45 | 9 (10.1) | 80 (89.9) |  |
|  | Oct-Dec-2024 | 22 | 11.1 | 3 (13.6) | 19 (86.4) |  |
| Current Fever | |  |  |  |  | 0.016 |
|  | No | 197 | 68.2 | 25 (12.7) | 172 (87.3) |  |
|  | Yes | 92 | 31.8 | 22 (23.9) | 70 (76.1) |  |
| Oropouche infection | |  |  |  |  |  |
|  | No | 67 | 23.2 | 0 (0.0) | 67 (100.0) | <0.001^b^ |
|  | Yes | 222 | 76.8 | 47 (100.0) | 175 (78.8) |  |

N, total number of participants

%, percentage

95% CI, 95% Confidence interval for incident cases.

^a^Chi-square test p value.

^b^Fisher Exact test p value.

**Table S5.** Poisson regression analysis of case contact study: bivariate and multivariable models adjusted for sex and age.

| Variables | Categories | RR | 95% CI | ^a^p-value | RR | 95% CI | ^b^p-value |
| --- | --- | --- | --- | --- | --- | --- | --- |
| Knows sick person outside home | | |  |  |  |  |  |
|  | No | Ref. |  |  |  |  |  |
|  | Yes | 1.05 | 0.89 - 1.24 | 0.542 |  |  |  |
| Sick Contact | |  |  |  |  |  |  |
|  | No | Ref. |  |  |  |  |  |
|  | Yes | 1.03 | 0.87 - 1.20 | 0.749 |  |  |  |
| Sick contact location | |  |  |  |  |  |  |
|  | At household | Ref. |  |  |  |  |  |
|  | Outside household | 1.08 | 0.90 - 1.28 | 0.411 |  |  |  |
|  | Both | 1.00 | 0.83 - 1.22 | 0.974 |  |  |  |
| Sick contact date | |  |  |  |  |  |  |
|  | February 2025 | Ref. |  |  | Ref. |  |  |
|  | January 2025 | 0.91 | 0.77 - 1.08 | 0.270 | 0.85 | 0.73 - 0.99 | 0.040 |
|  | Sep-Dec 2024 | 1.04 | 0.86 - 1.25 | 0.682 | 0.91 | 0.78 - 1.06 | 0.219 |
| Internal movement | |  |  |  |  |  |  |
|  | No | Ref. |  |  | Ref. |  |  |
|  | Yes | 0.98 | 0.85 - 1.11 | 0.710 | 0.98 | 0.87 - 1.12 | 0.816 |
| External movement | |  |  |  |  |  |  |
|  | No | Ref. |  |  |  |  |  |
|  | Yes | 0.98 | 0.86 - 1.12 | 0.775 |  |  |  |
| Fever in the last month | |  |  |  |  |  |  |
|  | No | Ref. |  |  |  |  |  |
|  | Yes | 1.72 | 1.39 - 2.12 | <0.001 |  |  |  |
| Fever Date |  |  |  |  |  |  |  |
|  | Feb-2025 | Ref. |  |  | Ref. |  |  |
|  | Jan-2025 | 1.02 | 0.91 - 1.14 | 0.716 | 1.14 | 0.98 - 1.32 | 0.083 |
|  | Oct-Dec-2024 | 0.93 | 0.76 - 1.14 | 0.506 | 0.96 | 0.74 - 1.26 | 0.789 |
| Current Fever | |  |  |  |  |  |  |
|  | No | Ref. |  |  |  |  |  |
|  | Yes | 1.31 | 1.17 - 1.46 | <0.001 |  |  |  |

^a^Bivariable Poisson regression (p value); adjusted for sex and age.

^b^Multivariable Poisson regression (p value); adjusted for sex and age.

RR, Relative risk

95% CI, 95% confidence interval

**Table S6**. List of all genomic data used in across the 4 datasets (L, M, S and concatenated segment datasets) (Excel)

**Table S7.** Selective pressure among codon sites for each OROV segment. (a) Fast unconstrained Bayesian approximation of pervasive selection (Posterior Probability > 0.90). (b) Site-specific episodic diversifying selection (Likelihood Ratio Test, LRT ≤ 0.05).

| Method | Large (L) segment | Medium (M) segment | Small (S) segment |
| --- | --- | --- | --- |
| FUBAR^a^ | 542 sites with evidence of  purifying selection. | 206 sites with evidence of  purifying selection | 9 sites with evidence of  purifying selection |
| MEME^b^ | 36 sites with evidence of episodic  diversifying selection | 25 sites with evidence of episodic  diversifying selection | 5 sites with evidence of episodic  diversifying selection |

**Table S8**. BF values for phylogeographic analyses within Latin America and Panama for the concatenated datasets.

|  | From | To | Bayes Factor |
| --- | --- | --- | --- |
| Latin America | Brazil | Colombia | Inf |
|  | Brazil | Bolivia | Inf |
|  | Cuba | Panama | 324.5 |
|  | Bolivia | Cuba | 60.5 |
|  | Peru | Brazil | 18.8 |
|  | French Guiana | Brazil | 7.8 |
| Panama | Meteti | Yaviza | 1406.7 |
|  | Meteti | Yape | 27.6 |
|  | Meteti | Lajas_Blanca | 23.7 |
|  | Meteti | Sambu | 7.3 |
|  | Meteti | Rio_Iglesia | 7.2 |
|  | Meteti | El_real | 6.9 |
|  | Meteti | Cemaco | 5.6 |

**Table S9.** Primer and probe sequences and final concentrations.

| **Name** | **Type** | **Sequence (5′ - 3′)** | **Genome Region** | **Evaluated concentration range** | **Final concentration in reaction** |
| --- | --- | --- | --- | --- | --- |
| OROV FS | Forward-S | GACAAGTSCTCAATGCTGGTGT | 92–113 | 0.1 µM - 0.9 µM | 0.4 µM |
| OROV FK | Forward-K | GACAAGTGCTCAATGCTKGTGT | 247–265 | 0.1 µM - 0.9 µM | 0.4 µM |
| OROV R | Reverse | CGTTGTCCGGSACTGGATT | 179–203 | 0.1 µM - 0.9 µM | 0.4 µM |
| OROV PY | Probe-Y | FAM- TGGTTGACCTYACTTTTGGTGGGGT -BHQ1 | N/A | 1.5 µM - 2.5 µM | 0.2 µM |
| OROV PR | Probe-R | FAM- TGGTTGACCTTACTTTTRGTGGGGT -BHQ1 | N/A | 1.5 µM - 2.5 µM | 0.2 µM |

**Table S10.** Data sources for risk modeling

| Type (Citation) | Dataset names | Description | Unit | Temporal  resolution | Spatial resolution |
| --- | --- | --- | --- | --- | --- |
| Soil features ^17^ | Soil Health Index (SHI) | Comprehensive index integrating physical, chemical and biological soil attributes to assess soil health across Latin America and Caribbean (LAC). | 1-5  1: least healthy  5: healthiest | Invariant | 90m |
|  | SHI soil function I | Storage and regulation of nutrient fluxes and availability. | Same as above | Invariant | 90m |
|  | SHI soil function II | Regulation of water fluxes, storage, and availability. | Same as above | Invariant | 90m |
|  | SHI soil function III | Soil organic carbon sequestration and biodiversity support. | Same as above | Invariant | 90m |
|  | SHI soil function IV | Physical support of plant growth. | Same as above | Invariant | 90m |
|  | SHI soil function V | Resistance to erosion and degradation. | Same as above | Invariant | 90m |
| Weather ^18^ | 2m temperature-mean | 24-hour mean of the air temperature measured at a height of 2 meters above the Earth’s surface. | K | Daily | 0.1° × 0.1° (~11 km) |
|  | 2m temperature-min | 24-hour minimum of the air temperature measured at a height of 2 meters above the Earth’s surface. | K | Daily | 0.1° × 0.1° (~11 km) |
|  | 2m temperature-max | 24-hour maximum of the air temperature measured at a height of 2 meters above the Earth’s surface. | K | Daily | 0.1° × 0.1° (~11 km) |
|  | Precipitation flux | The rate at which precipitation (rain, snow, etc.) falls to the ground over a specific area and time period. | mm per day | Daily | 0.1° × 0.1° (~11 km) |
|  | Vapor pressure | The contribution of water vapor to the total atmospheric pressure over 24 hours (00h-24h) in local time. | hPa | Daily | 0.1° × 0.1° (~11 km) |
| Climate ^10^ | Temperature-monthly^4^ | The mean, minimum, and maximum of the monthly temperature. | C | Monthly | 0.5′ × 0.5′  (~0.9 km) |
|  | Precipitation-monthly^4^ | The monthly precipitation. | mm | Monthly | 0.5′ × 0.5′  (~0.9 km) |
|  | Vapor pressure-monthly^4^ | The contribution of water vapor to the total atmospheric pressure over a month in local time. | kPa | Monthly | 0.5′ × 0.5′  (~0.9 km) |
| Population ^19,20^ | Human footprint^6^ | A quantitative measure of human influence on the nature environment. | N/A | Invariant | N/A |
|  | Global population density^7^ | The number of people living per unit area of land across the Earth’s surface. | Persons/km^2^ | Every 5 years | 30 arcsecond |
| Agriculture^21,22^ | CROPGRIDS | The amount of land dedicated to production of specific crops (banana, cassava, cocoa, coffee, rubber, soybean, sugarcane) | Hectares/area | Invariant | 0.05° × 0.05° (~5.6 km) |
|  | FAO Categories | FAO categorization of specific crops (e.g., cereals, sub/tropical fruits, etc.); used to group specific crops by type | N/A | N/A | N/A |

**Table S11.** Model AICs

| Model Stage | Model AIC | ∆ AIC |
| --- | --- | --- |
| Fit using top variables identified by Random Forest | 1078.046 |  |
| Fit Following Step Reduction | 1063.521 | -14.5256 |
| Minimum Adequate Model (Final) | 1060.300 | -3.220 |

**Table S12.** Manual Removal Steps and AIC values

| Removed Variables | AIC | ∆ AIC |
| --- | --- | --- |
| Fit Following Step Reduction (No removals) | 1063.521 |  |
| Mean Daily Average Temperature In Subsequent 30 Days | 1063.821 | 0.300 |
| Mean Daily Low Temperature In Subsequent 30 Days | 1062.321 | -1.499 |
| Mean Daily Average Temperature In Previous 30 Days | 1061.228 | -1.093 |
| Mean Daily High Temperature In Previous 30 Days | 1059.859 | -1.369 |
| Percent Beverage Crops (FAO) | 1059.976 | 0.117 |
| Soil Water Regulation Ability | 1060.300 | 0.325 |

**Table S13.** Coefficients and Estimates from Minimum Model (fit to normalized Brazilian data)

| Variable | Estimate | Std Error | z value | p-value |
| --- | --- | --- | --- | --- |
| Intercept | -2.233 | 0.281 | -7.947 | <0.0001 |
| Mean Daily Precipitation In Subsequent 30 Days | 0.583 | 0.142 | 4.090 | <0.0001 |
| Precipitation In Month With Most Precipitation In A Typical Year | -0.511 | 0.084 | -6.234 | 0.0000000 |
| Percent Sub/Tropical Fruit (FAO) | 0.263 | 0.064 | 4.047 | 0.0000520 |
| Mean Daily Average Vapor Pressure in Previous 30 Days | 1.201 | 0.3040 | 3.949 | 0.0000783 |
| Mean Daily High Temperature In Subsequent 30 Days | 0.972 | 0.244 | 3.983 | 0.0000681 |
| Log 10 of Density-Weighted Population Density | -0.480 | 0.107 | -4.479 | 0.0000075 |
| Percent Coffee Cultivation | 0.234 | 0.075 | 3.130 | 0.0017467 |
| Mean Daily Low Temperature In Previous 30 Days | -1.134 | 0.446 | -2.5442 | 0.0109521 |
| Maximum Human Footprint | 0.468 | 0.098 | 4.772 | 0.0000018 |
| Precipitation In Month With Least Precipitation In A Typical Year | 0.262 | 0.098 | 2.664 | 0.0077131 |
| Percent Cereal Crops (FAO) | -1.393 | 0.345 | -4.0405 | 0.0000533 |

**Table S14.** Laboratory confirmation of cases by surveillance component and diagnostic approach.

| **Surveillance components** | **Total Enrolled** | **Tested Negative** | **PCR** | **IgM** | **IgG/PRNT** | **IgM/IgG** | **PCR/IgM** | **Total Positive** |
| --- | --- | --- | --- | --- | --- | --- | --- | --- |
| Health Care Centers | 690 | 476 | 148 | 64 | 0 | 2 | 0 | 214 |
|  |  |  | 1 (0–2)* | 10.5 (7–16)* |  |  |  |  |
| Field epidemiological investigation (With symptoms) | 156 | 52 | 9 | 79 | 3 | 11 | 2 | 104 |
|  |  |  | 1 (0–2)* | 7 (2–22)* | 12.5 (5–31.5)* |  |  |  |
| Field epidemiological investigation (Without symptoms) | 136 | 63 | 3 | 70 | 0 | 0 | 0 | 73 |
| Contact-based telephone (With symptoms) | 58 | 2 | 55 | 1 | 0 | 0 | 0 | 56 |
|  |  |  | 1 (0–2)* | 74 |  |  |  |  |
| Contact-based telephone (Without symptoms) | 0 | 0 | 0 | 0 | 0 | 0 | 0 | 0 |
| **Pooled summary across surveillance components** | **1040** | **593** | **215** | **214** | **3** | **13** | **2** | **447** |
|  |  |  | 1 (0–2)* | 10 (5–19)* | 12.5 (5–31.5)* |  |  |  |

* Median days refer to days after symptom onset at the time of sample collection. IQR: interquartile range.

Cases identified among asymptomatic participants do not have symptom-onset timing available.

“Total Positive” corresponds to laboratory-confirmed OROV infections/recent exposures identified by PCR, ELISA IgM, ELISA IgG/PRNT, combined IgM/IgG ELISA, or combined PCR/IgM positivity.

**Table S15.** OROV cases by corregimiento and district, Darién Province, Panama

| District | Corregimiento | OROV Cases (n) |
| --- | --- | --- |
| Pinogana | Metetí | 170 |
| Pinogana | Yaviza | 47 |
| Pinogana | El Real de Santa María | 21 |
| Pinogana | Yape | 9 |
| Pinogana | Comarca Kuna de Wargandí | 1 |
| Santa Fe | Río Iglesias | 46 |
| Santa Fe | Santa Fe | 13 |
| Santa Fe | Agua Fría | 5 |
| Santa Fe | Río Congo | 5 |
| Santa Fe | Zapallal | 5 |
| Santa Fe | Cucunatí | 2 |
| Santa Fe | Río Congo Arriba | 2 |
| Santa Fe | Not specified | 3 |
| Chepigana | Garachiné | 20 |
| Chepigana | La Palma | 8 |
| Chepigana | Camogantí | 4 |
| Chepigana | Jaqué | 4 |
| Chepigana | Taimatí | 5 |
| Cémaco | Lajas Blancas | 29 |
| Cémaco | Manuel Ortega | 4 |
| Sambú | Sambú | 4 |
| Sambú | Río Sábalo | 1 |

**Table S16. Demographics and characteristics of participants with samples analized for viral load and cytokine response.**

| Characteristic | All participants (N=97) | OROV acute samples (≤ 7 days of symptoms onset) (N=56) | OROV negative healthy individuals (N=41) | P-value* |
| --- | --- | --- | --- | --- |
| Age (years) |  |  |  | P=0.94 |
| mean | 32 | 33 | 32 |  |
| range | 16 - 79 | 16 - 79 | 17 - 72 |  |
| Sex, n (%) |  |  |  | P=0.87 |
| Female | 55 (56.7%) | 33 (58.9%) | 22 (53.7%) |  |
| Male | 42 (43.3%) | 23 (41.1%) | 19 (46.3%) |  |
| Days of symptoms onset |  |  |  |  |
| mean | NA | 1 | NA |  |
| range | NA | 0-7 | NA |  |

*P value indicates if differences between groups are statistically significant (<0.05). Mann Whitney U test was used for age and Chi square for gender.

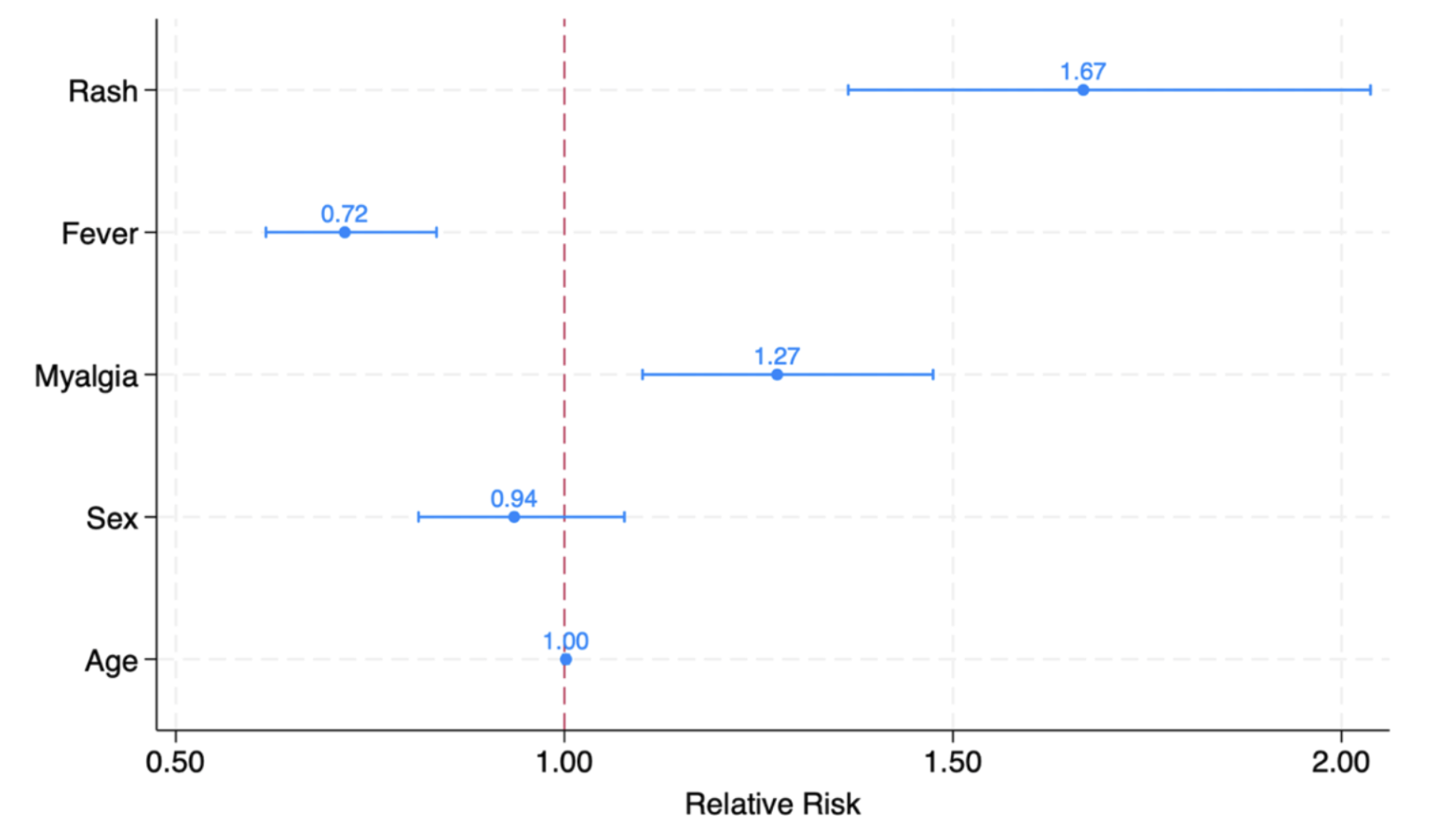

**Figure S1**. Symptoms associated with OROV infection in the final multivariable Poisson model. Relative risk (RR) and 95% confidence intervals for variables selected through forward stepwise nested modelling. The model includes age and sex as adjustment variables. The dashed red line denotes the no-effect threshold (RR = 1.0).

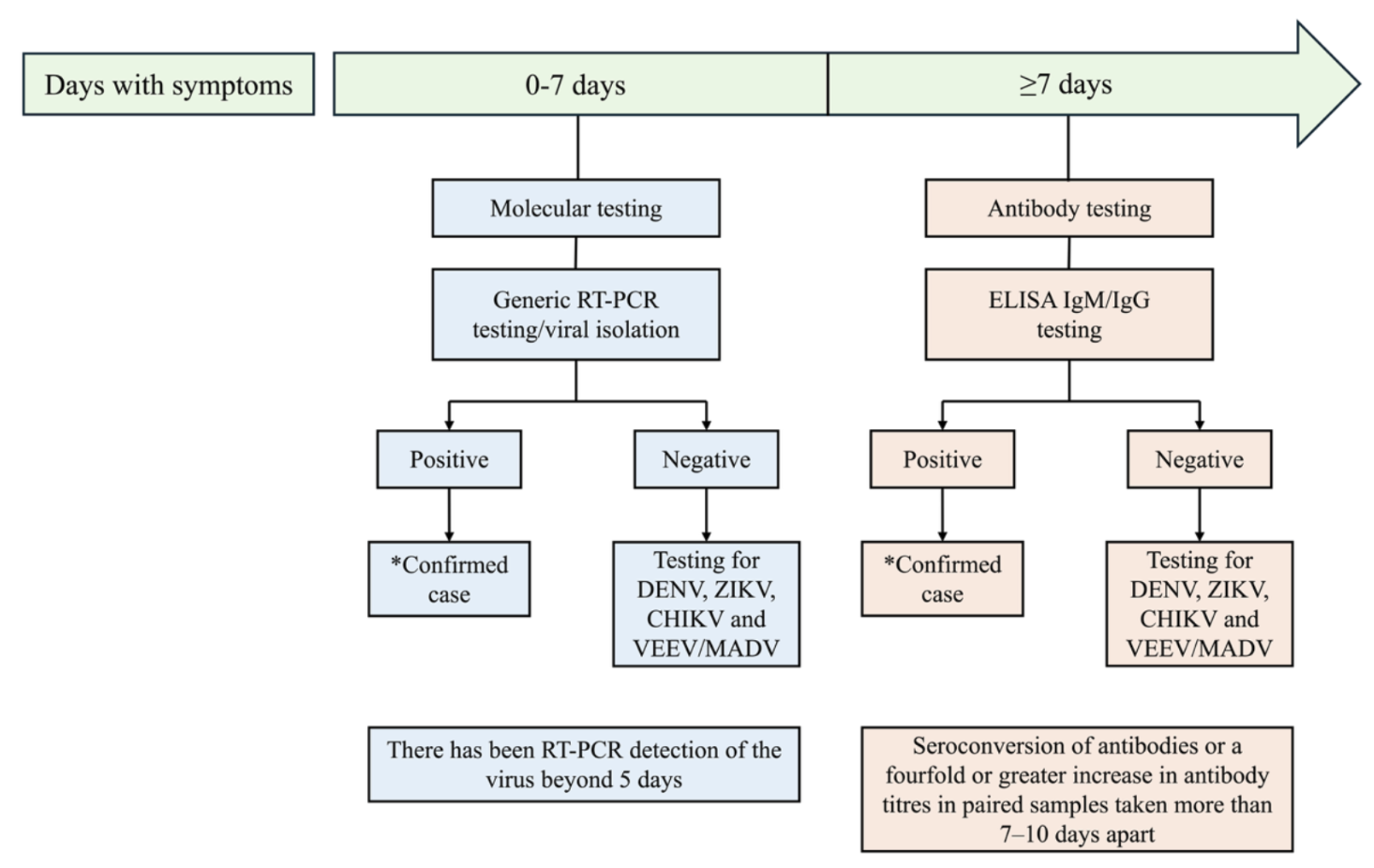

**Figure S2.** General laboratory algorithm used for diagnosis of OROV infections

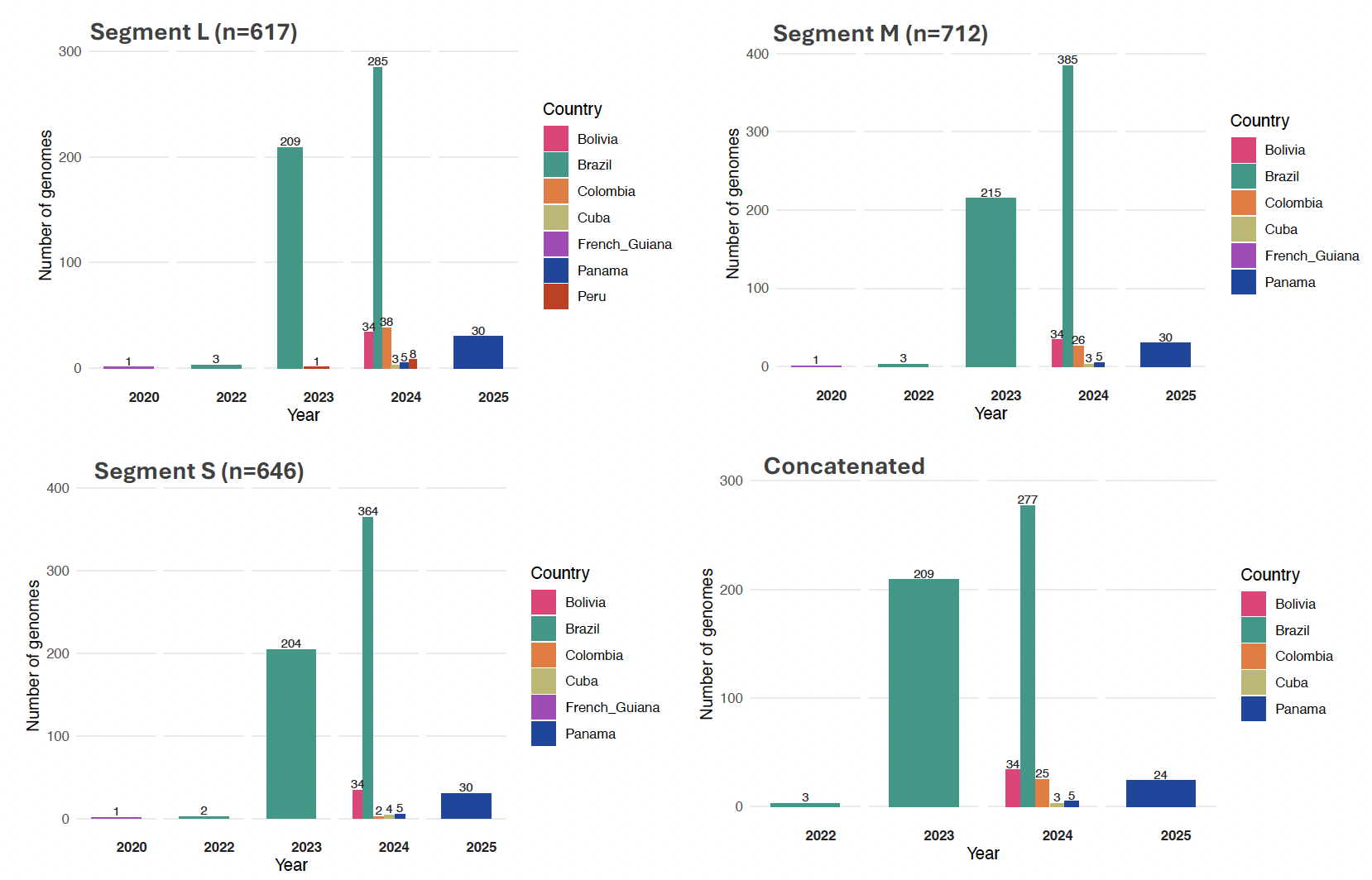
**Figure S3.** Temporal distribution of OROV sequences available in public data bases and generated for this study, stratified by genomic segments (L, M, S, and concatenated), from 2020 to 2025 in the study region. This image illustrates the trends in sequencing efforts and data availability across time and viral segments.

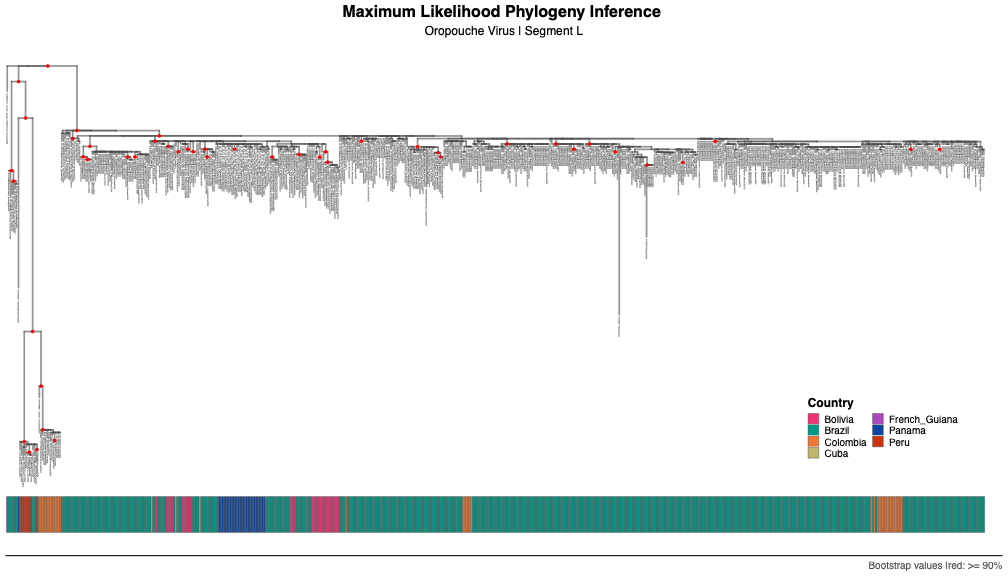

**Figure S4.** Maximum likelihood phylogenies of 612 non-recombinant OROV L segment. Red circles indicate strong statistical support along the branches defined by bootstrap (BS) values ≥ 90.

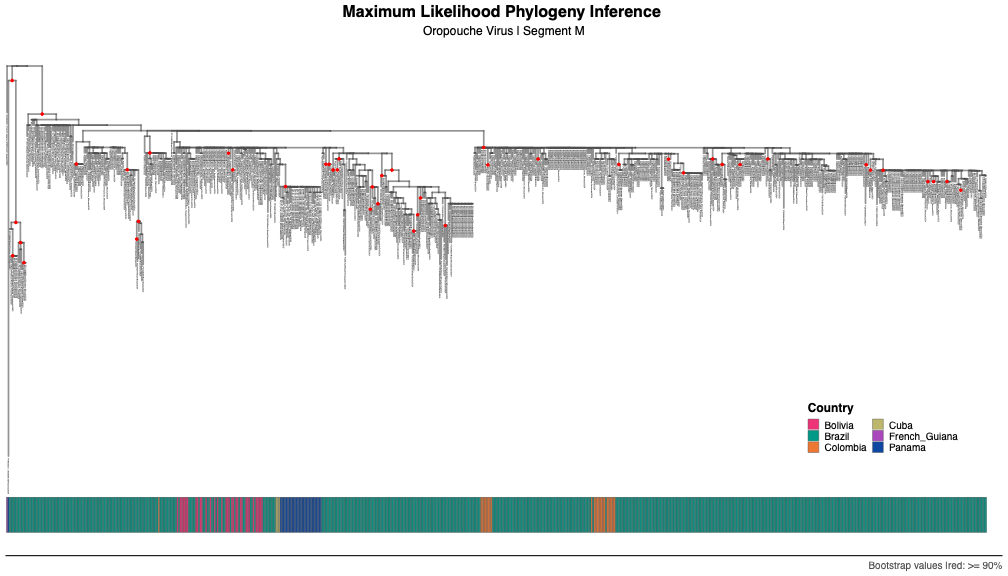

**Figure S5.** Maximum likelihood phylogenies of 690 non-recombinant OROV M segment. Red circles indicate strong statistical support along the branches defined by bootstrap (BS) values ≥ 90.

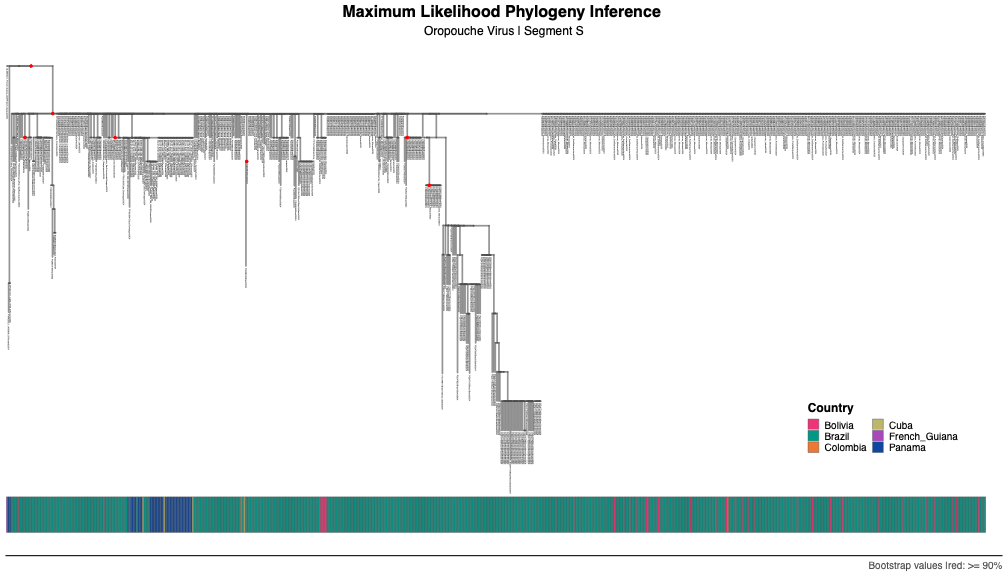

**Figure S6.** Maximum likelihood phylogenies of 646 non-recombinant OROV S segment. Red circles indicate strong statistical support along the branches defined by bootstrap (BS) values ≥ 90.

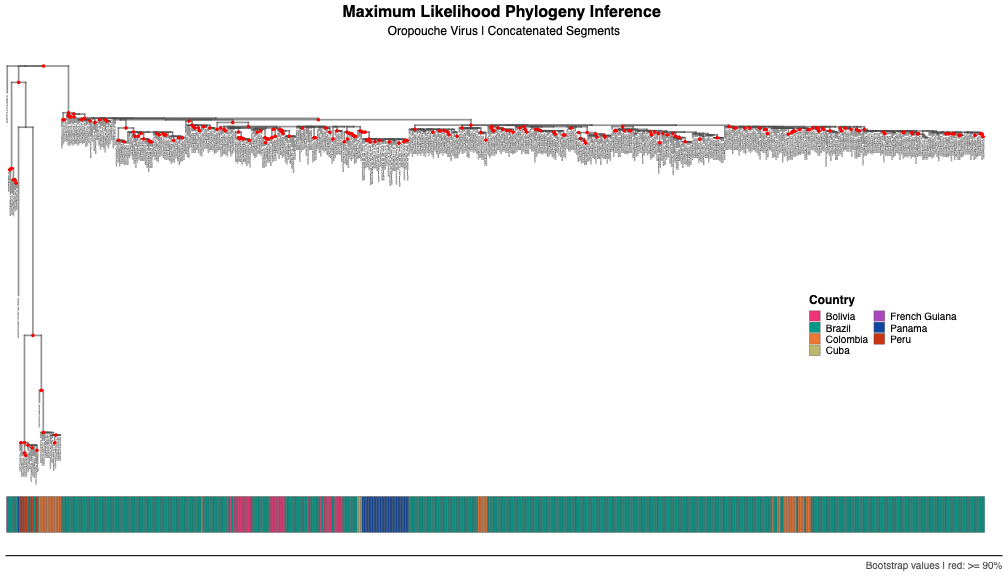

**Figure S7**. Maximum likelihood phylogenies of 607 non-recombinant OROV genomic concatenated segments. Red circles indicate strong statistical support along the branches defined by bootstrap (BS) values ≥ 90.

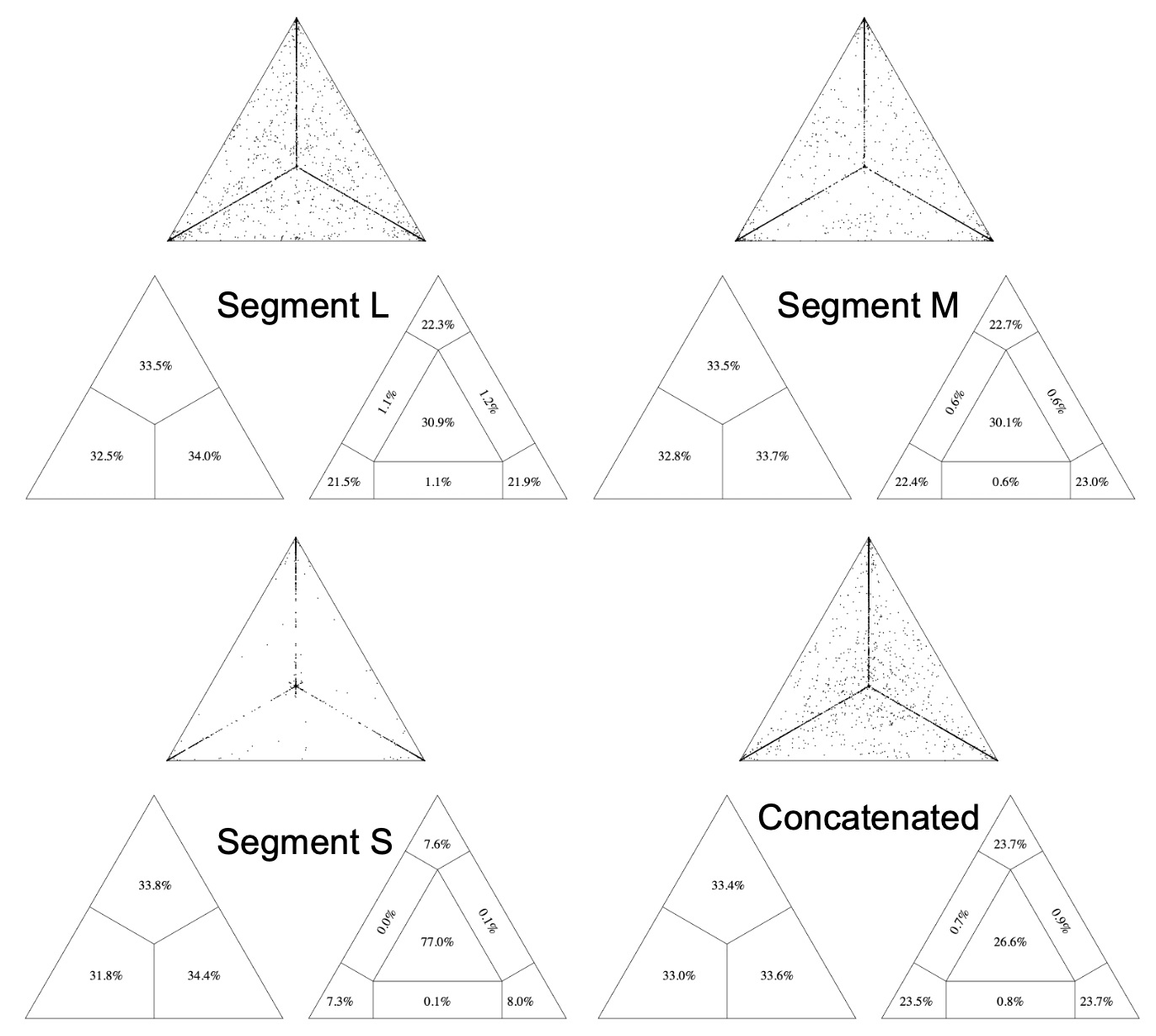

**Figure S8.** Phylogenetic signal of OROV maximum likelihood phylogenies. This analysis was stratified by genomic segments (L, M, S, and concatenated) and reflects trees in figures S3, S4, S5 and S6.

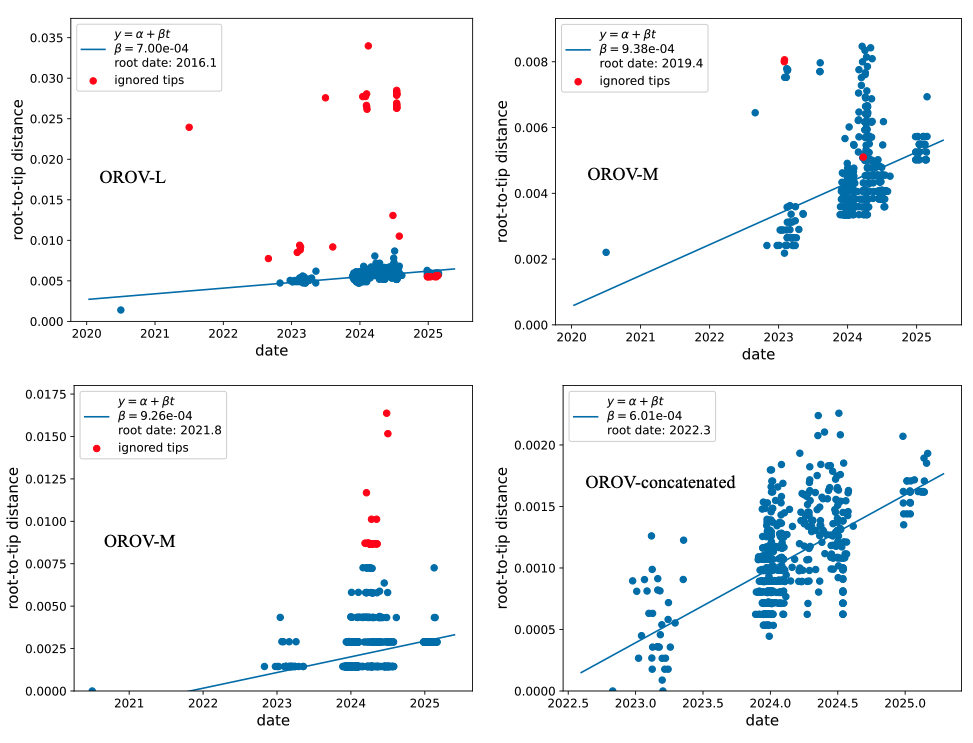

**Figure S9.** Temporal signal of OROV genomic segments L, M, S, and concatenated. Root-to-tip linear regression between sampling date and genetic distance, based on best-fitting root based on trees in figures S3, S4, S5 and S6, estimated using a heuristic residual mean squared optimization approach.

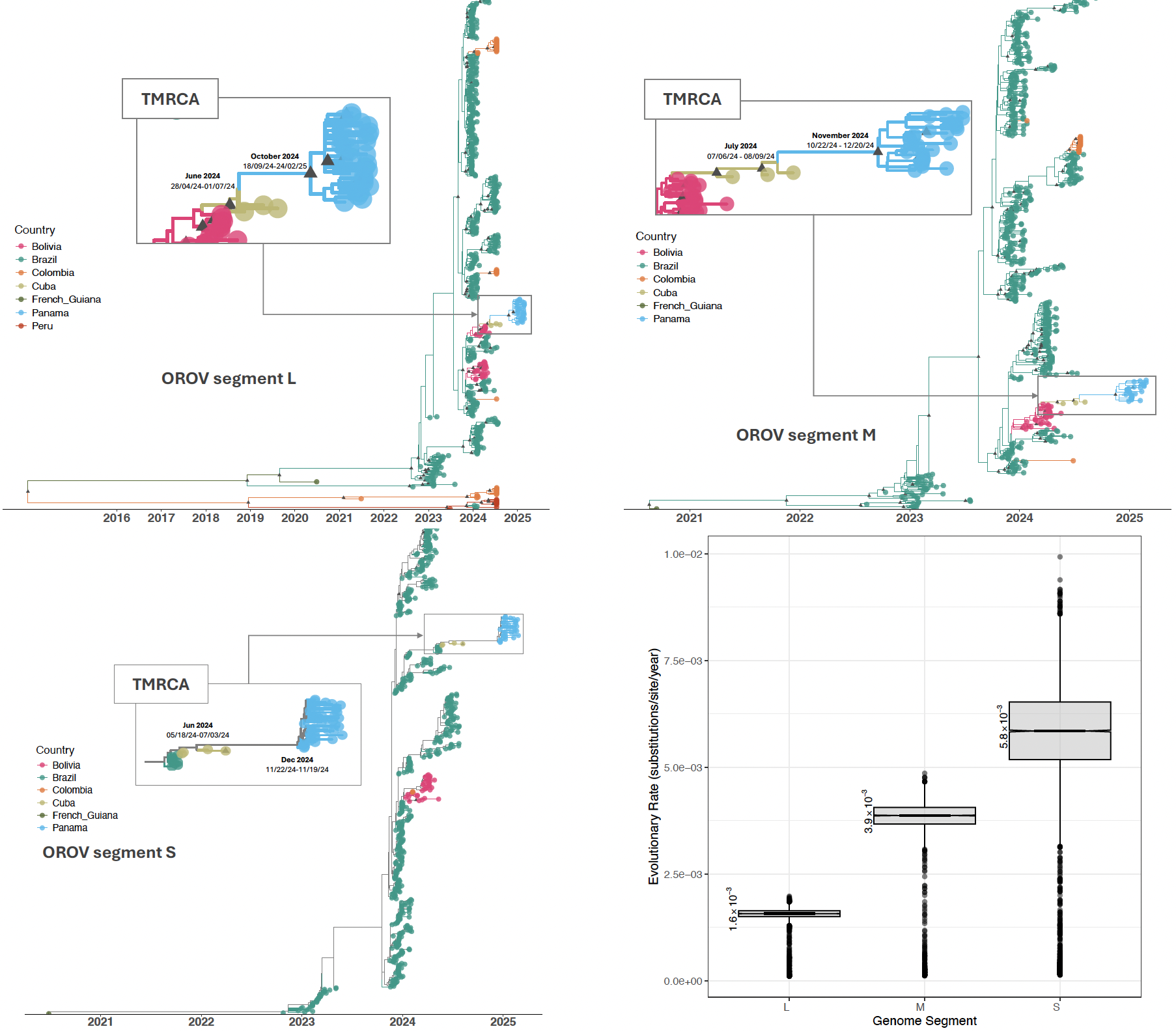

**Figure S10.** Spatial dissemination of OROV L (n=608), M (n=684), and S (n=638) full-genome sequenced samples from 2020 to 2025 confirmed cases in Latin America. Bayesian scale MCC tree under a discrete phylogeographic model. Branches and tips are colored according to the most probable ancestral location, with a posterior probability (PP)≥0.90 (black triangle). The accompanying map shows the spatial spread of OROV among eight sampled locations, with transition routes in the region supported by Bayes Factor (BF) evidence (strong support: BF ≥100; moderate support 5 ≤ BF < 100). The boxplot shows the evolutionary rate with the molecular clock median value and 95% HPD estimated by Bayesian phylogeography for each viral segment, using samples obtained from 2020 to 2025 confirmed cases in Latin America.

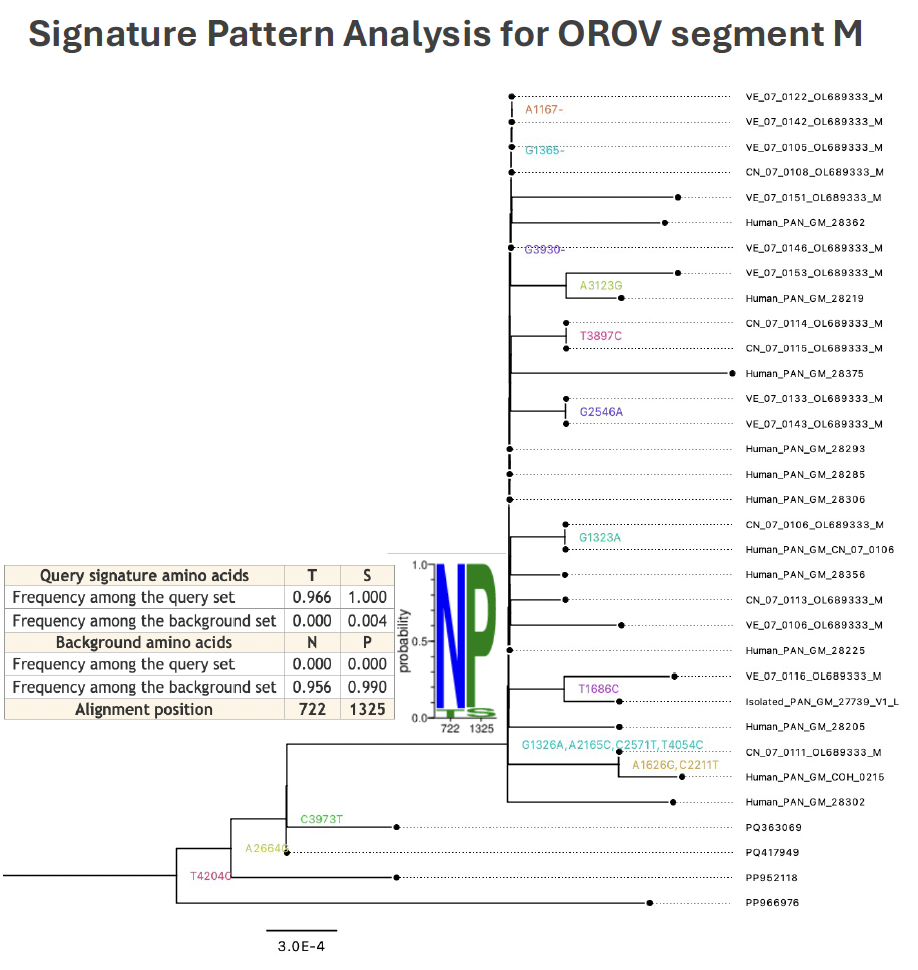

**Figure S11.** Signature pattern analysis for OROV M segment. We assessed viral sequence relatedness from amino acids sequences between OROV_PAN2024-2025_ clade and background sequences using the dataset reported in the tree in figure S2. The tree is a zoom from the phylogenetic maximum likelihood tree of the OROV_PAN2024-2025_ clade in figure S4 obtained from Treetime, using ancestral reconstruction methods including the amino acid data. T722N and S1325P were found in M segment of OROVPAN2024-2025, however were found to be neutral based on the following cut-offs: LRT p ≤ 0.05 for aBSREL and posterior probability (PP) > 0.90 for FUBAR, and LRT ≤ 0.05 for MEME.

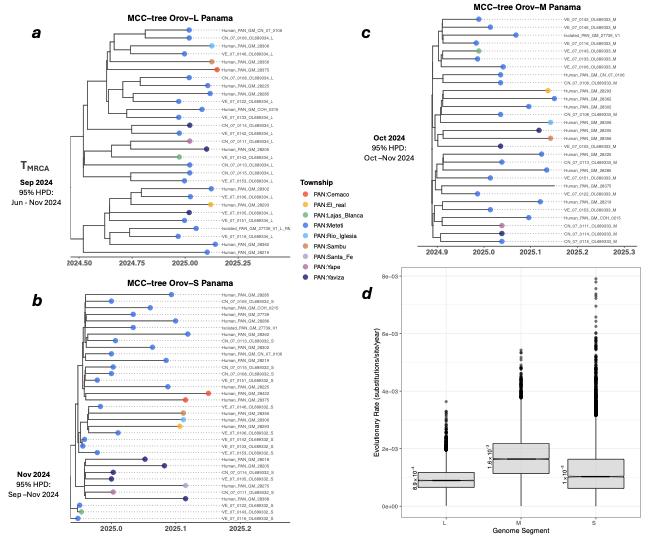

**Figure S12.** Temporal dynamics of the current OROV_PAN2024-2025_ clade across the Darién and Emberá provinces, Panama. MCC inferred from each of the genomic viral segments (L, M, S – concatenated in main manuscript). The boxplot shows the evolutionary rate with the molecular clock median value and 95% HPD for each viral segment and concatenated.

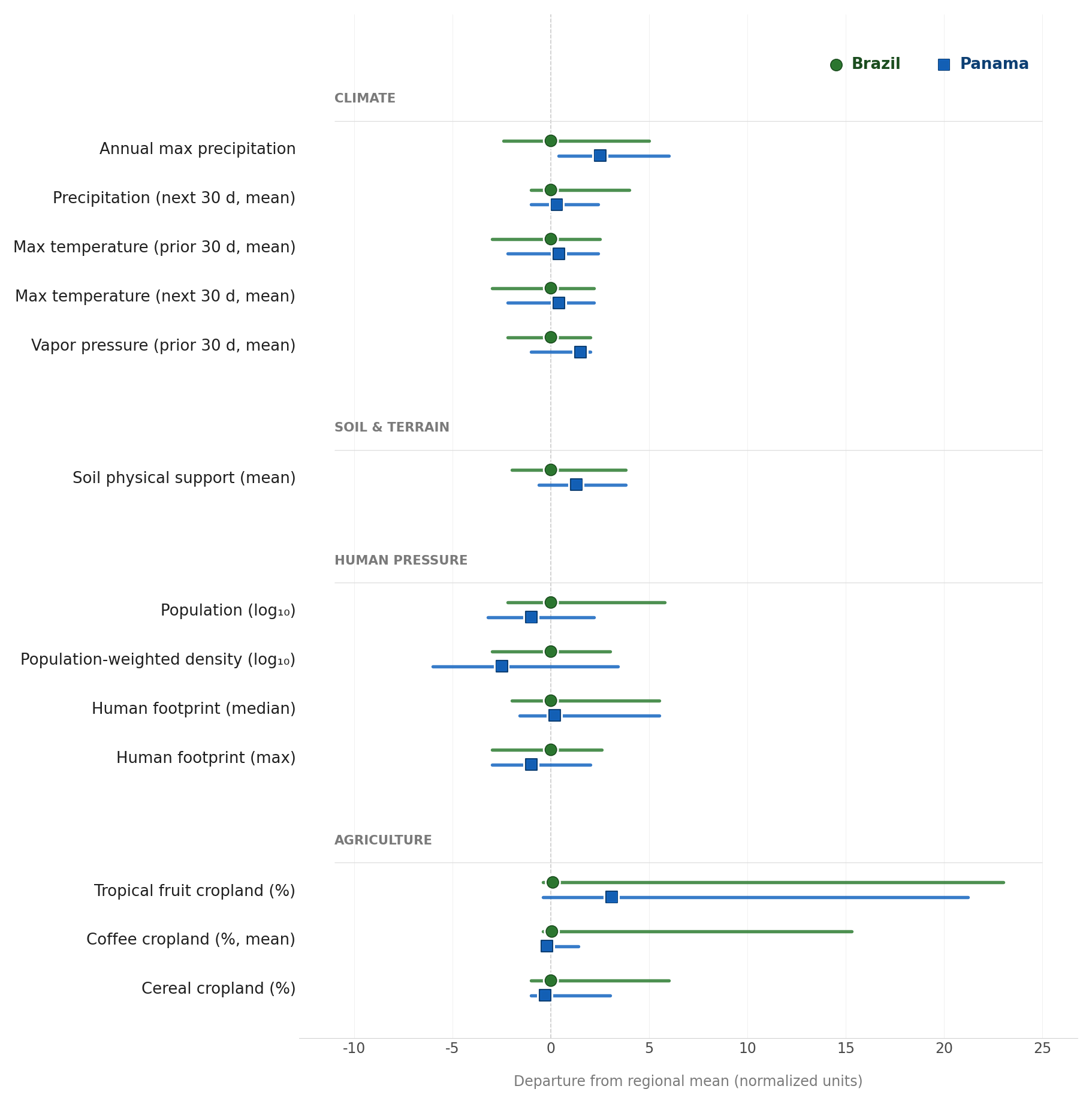

**Figure S13.** Range and mean values for normalized predictors. Mean value indicated by points. Predictors’ ranged and mean values are indicated for Brazil (green) and Panama (blue).

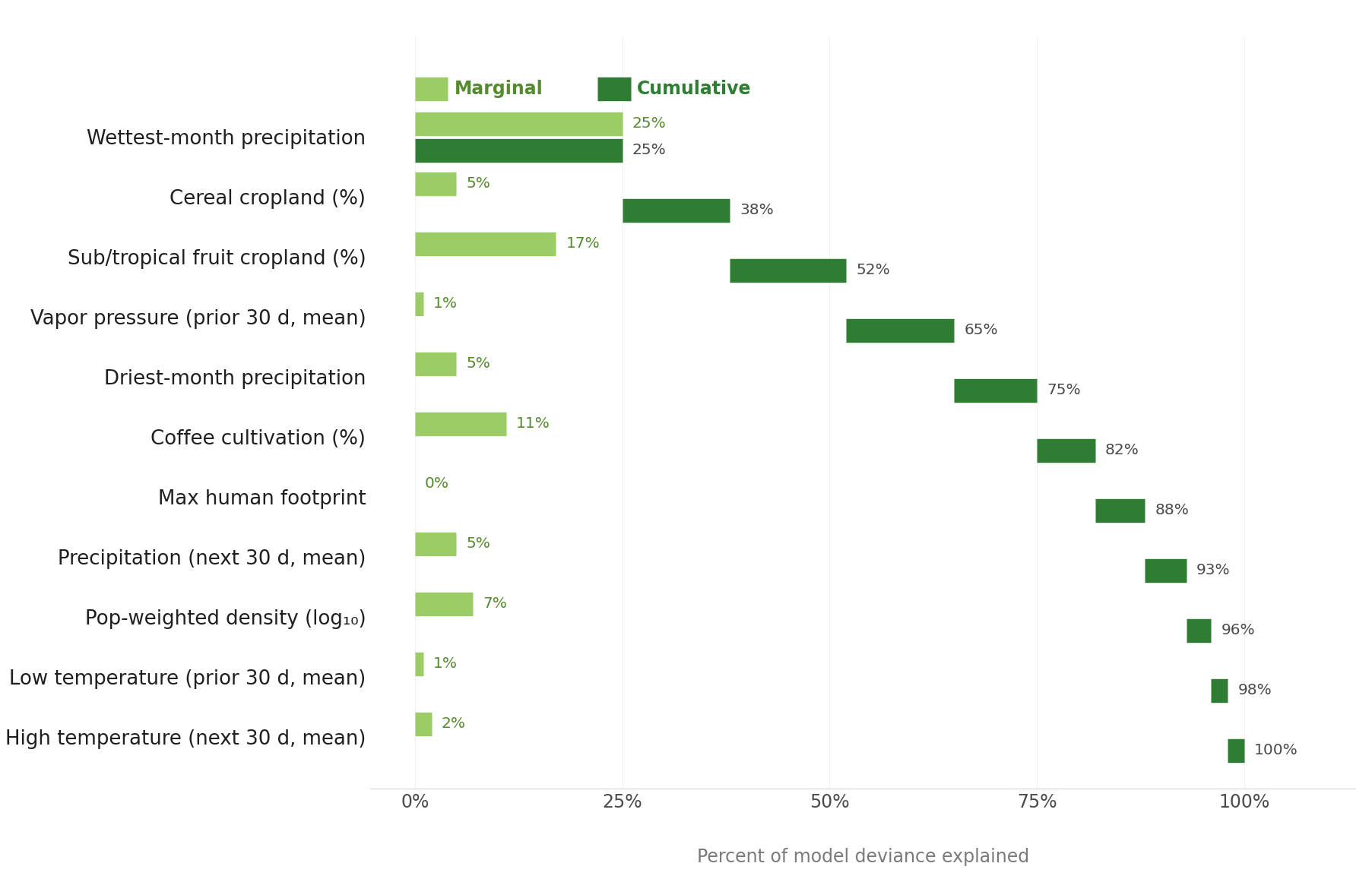

**Figure S14.** Variable importance plot for minimum adequate model, fit to Brazilian data. Marginal (light green) and cumulative (dark green) deviance explained by each explanatory variable. Refer to Table S13 for sign, magnitude, and statistical significance of each effect.

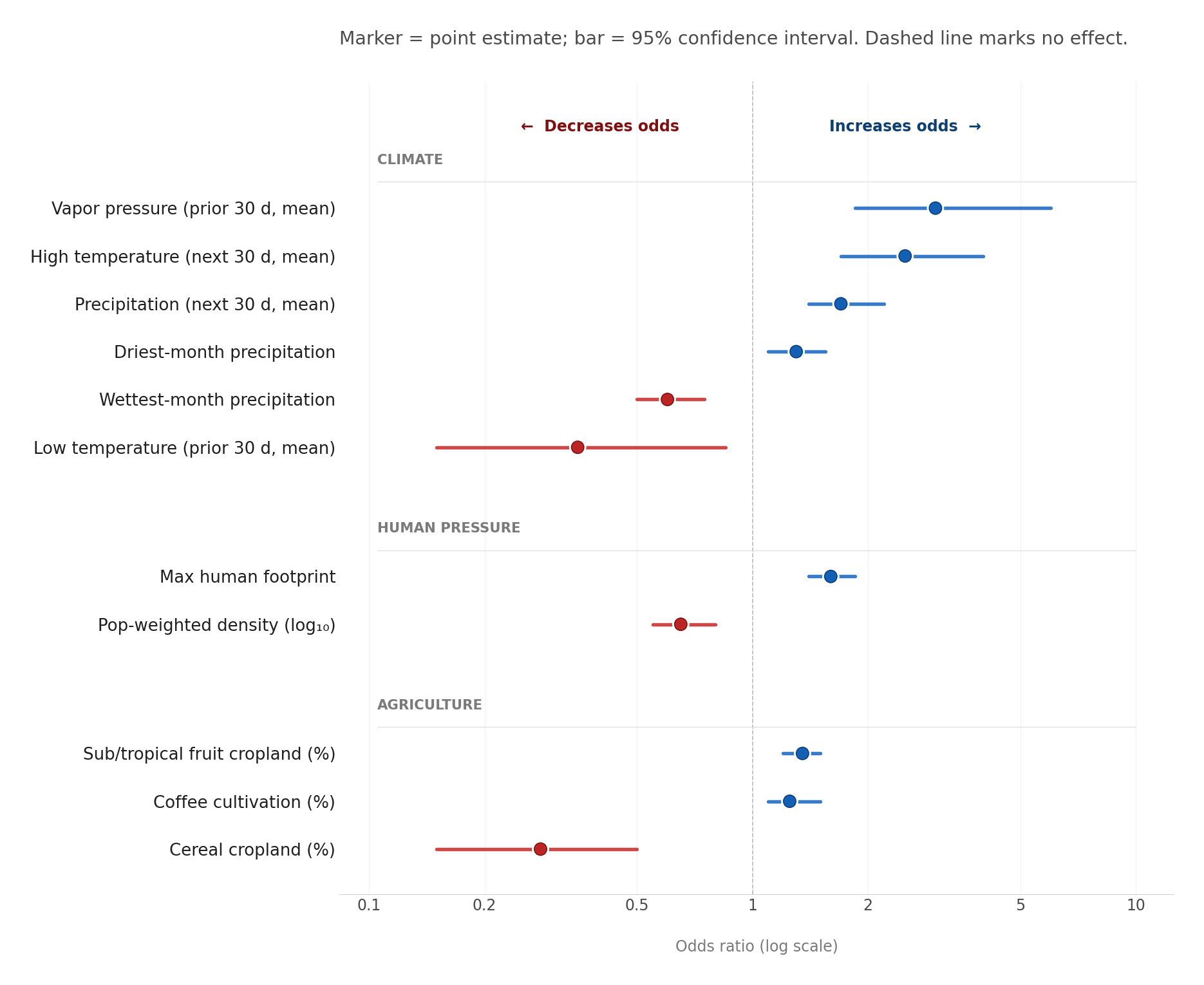

**Figure S15.** Odds Ratios and 95% confidence intervals for the 11 predictors in the OROV outbreak probability regression model. Dots represent the odds ratio, and bars indicate the 95% confidence intervals. Blue indicates an increase in outbreak risk with an increase in the predictor value; red indicates a decrease in outbreak risk with an increase in the predictor value.

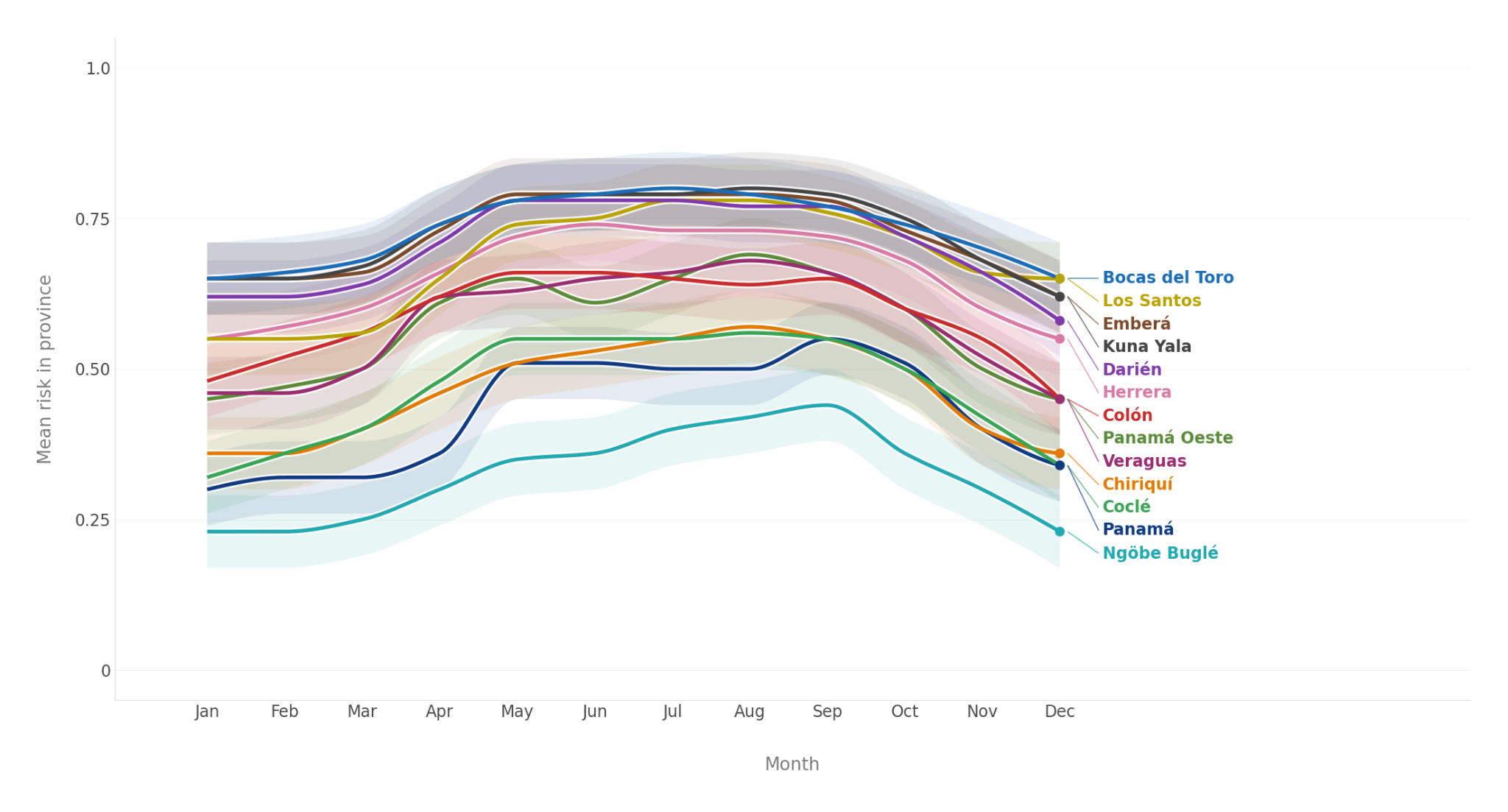

**Figure S16.** Variation in mean risk through the year by province, projecting from model fit to Brazilian data. Each line represents the typical monthly risk for one Panamanian province, as estimated from climate normals; shaded bands show 95% conf. intervals. All provinces share a similar unimodal seasonal peak in the wet season, but baseline risk varies substantially, from year-round high values in Bocas del Toro, Kuna Yala, Emberá and Darién, to consistently lower values in Ngöbe Buglé and Panamá. No corregimientos were significantly out of phase.

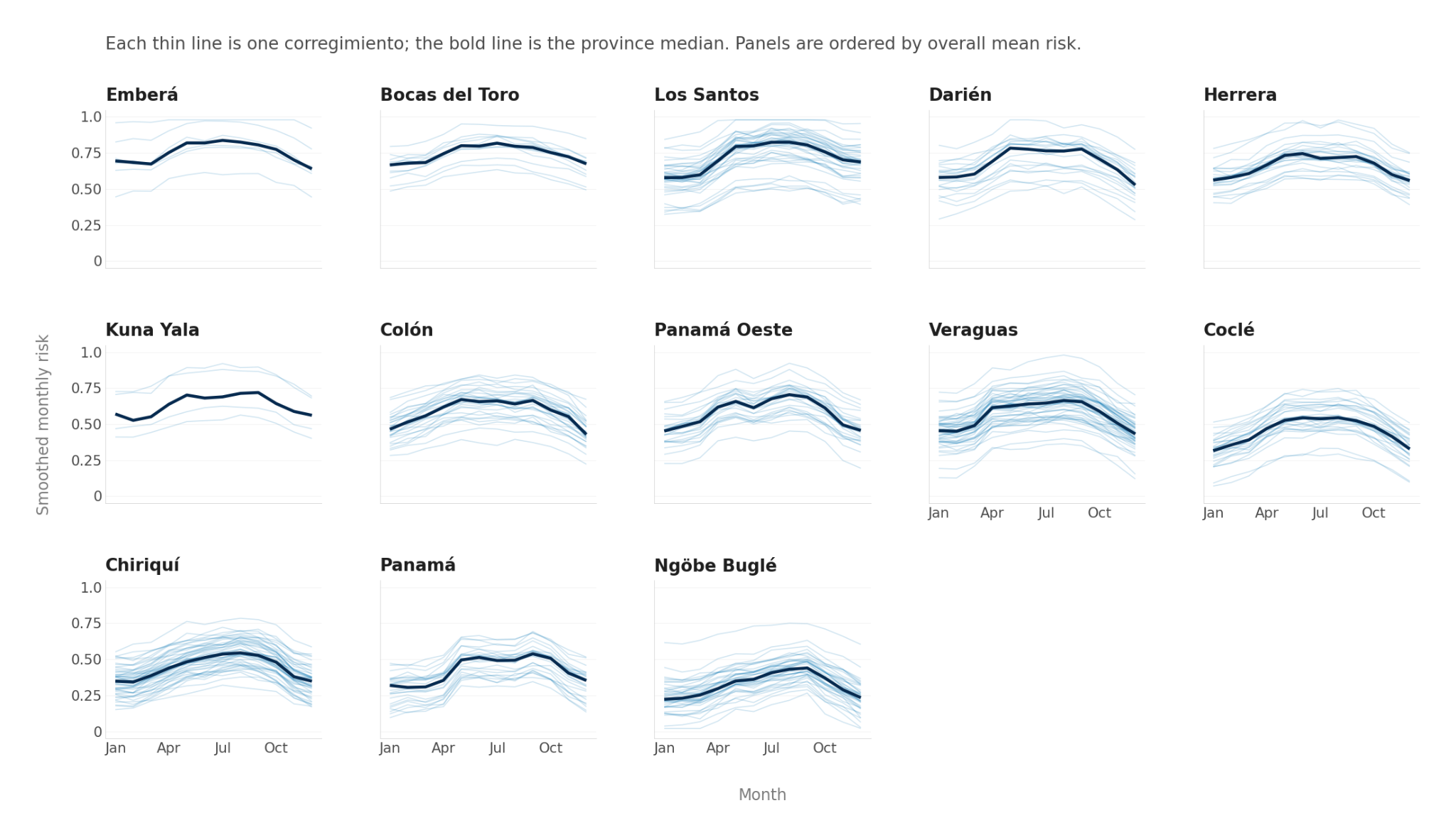

**Figure S17.** Variation in risk through the year, projecting from model fit to Brazilian data. Each column represents a province, as labeled. Each line represents the typical risk by month for one corregimiento, as estimated from climate normals. No corregimientos were significantly out of phase. The bold line shows the province median.

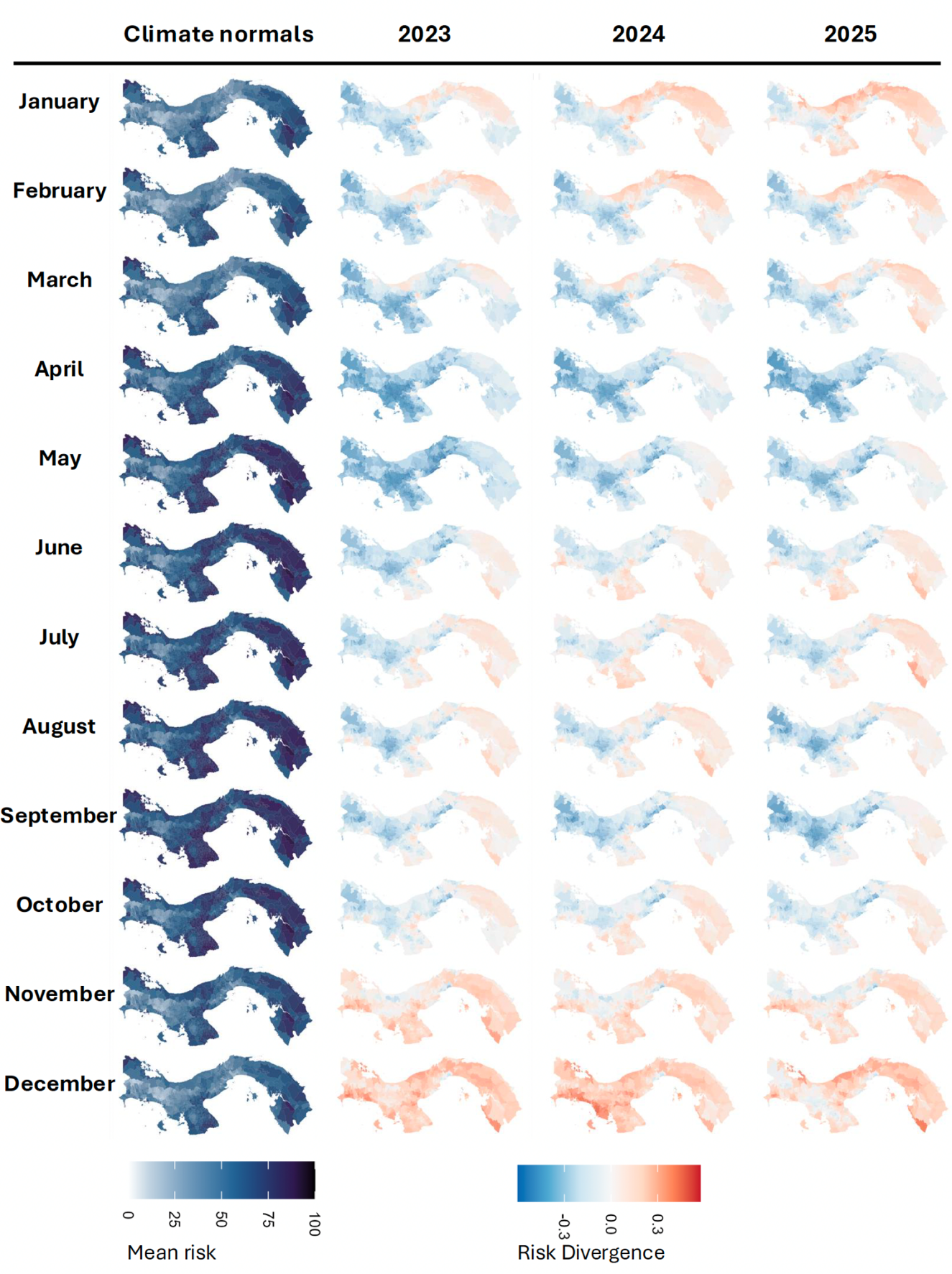

**Figure S18.** Typical risk estimates and divergence from typical risk by month. Leftmost column: Predicted risk by month, using climate normals for expected weather for each month. Columns to the right: Risk divergence (risk calculated from observed weather minus risk predicted from expected weather in that month based on climate normals) for 2023, 2024, and 2025.

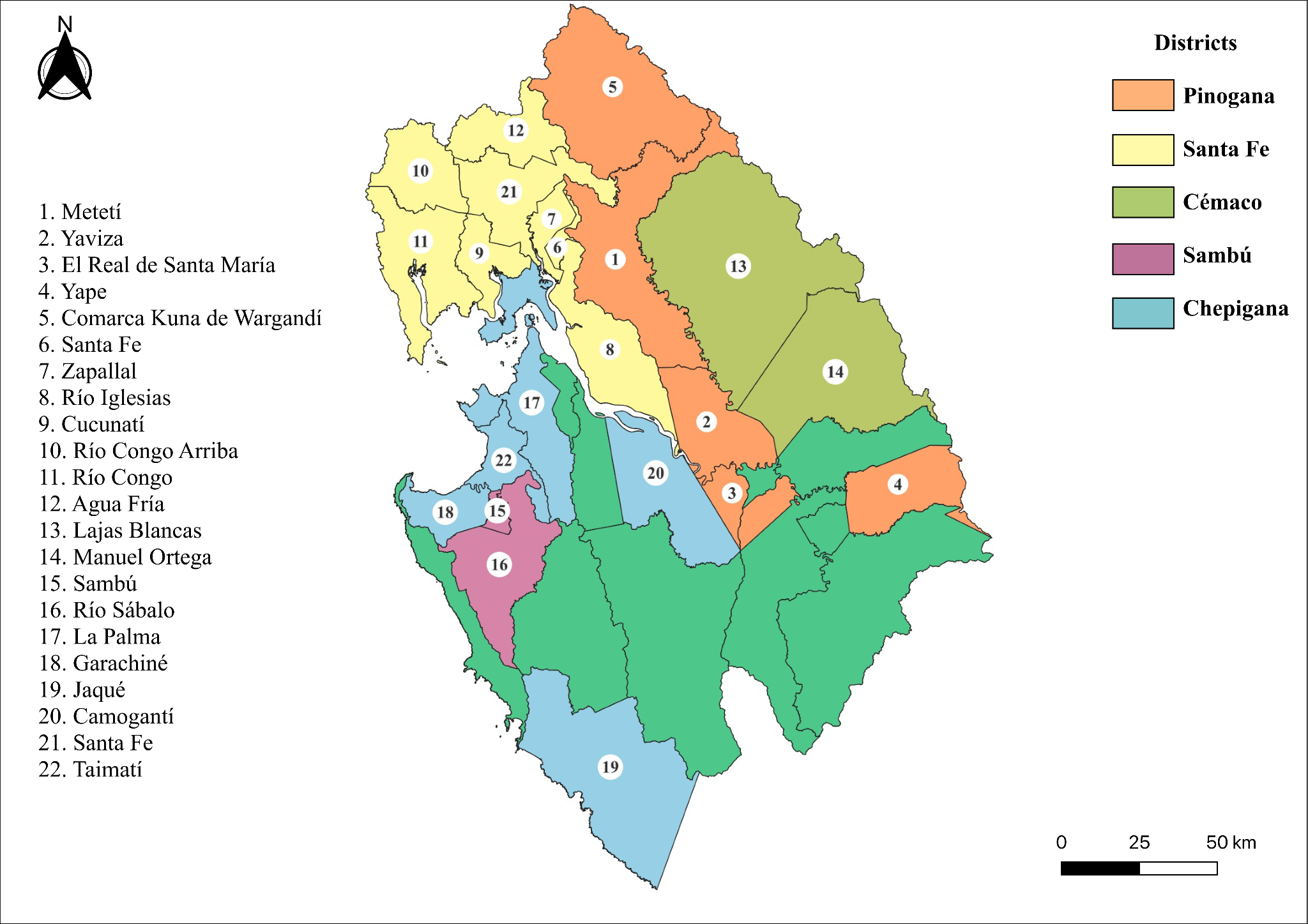

**Figure S19.** Geographical distribution of OROV cases in Darién Province, Panama. Map of Darién Province showing locations where OROV cases were identified during the study period. Colours indicate administrative districts (Pinogana, Santa Fe, Cémaco, Sambú, and Chepigana). Numbered markers denote corregimientos as indicated in the figure.

**References**

1.	Landis, J. R. & Koch, G. G. The measurement of observer agreement for categorical data. *Biometrics* **33**, 159–174 (1977).

2.	Ma, J. & Earn, D. J. D. Generality of the Final Size Formula for an Epidemic of a Newly Invading Infectious Disease. *Bull. Math. Biol.* **68**, 679–702 (2006).

3.	Brown, L. D., Cai, T. T. & DasGupta, A. Interval Estimation for a Binomial Proportion. *Statist. Sci.* **16**, (2001).

4.	Hijmans, R. J., Barbosa, M., Ghosh, A. & Mandel, A. *Geodata: Download Geographic Data*. (2025).

5.	Daniel Baston. exactextractr: Fast Extraction from Raster Datasets using Polygons. 0.10.0 https://doi.org/10.32614/CRAN.package.exactextractr (2019).

6.	Pereira, R. H. M. & Goncalves, C. N. *Geobr: Download Official Spatial Data Sets of Brazil*. (2025).

7.	Kamvar, Z. N., Cai, J., Pulliam, J. R. C., Schumacher, J. & Jombart, T. Epidemic curves made easy using the R package *incidence*. Preprint at https://doi.org/10.12688/f1000research.18002.1 (2019).

8.	Ishwaran, H. & Kogalur, U. B. *Fast Unified Random Forests for Survival, Regression, and Classification (RF-SRC)*. (manual, 2025).

9.	R Core Team and contributors worldwide. R: The R Stats Package. (2025).

10.	Fick, S. E. & Hijmans, R. J. WorldClim 2: new 1‐km spatial resolution climate surfaces for global land areas. *Intl Journal of Climatology* **37**, 4302–4315 (2017).

11.	Murrell, B. *et al.* Detecting Individual Sites Subject to Episodic Diversifying Selection. *PLoS Genet* **8**, e1002764 (2012).

12.	Murrell, B. *et al.* FUBAR: A Fast, Unconstrained Bayesian AppRoximation for Inferring Selection. *Molecular Biology and Evolution* **30**, 1196–1205 (2013).

13.	Smith, M. D. *et al.* Less Is More: An Adaptive Branch-Site Random Effects Model for Efficient Detection of Episodic Diversifying Selection. *Molecular Biology and Evolution* **32**, 1342–1353 (2015).

14.	Korber, B. & Myers, G. Signature Pattern Analysis: A Method for Assessing Viral Sequence Relatedness. *AIDS Research and Human Retroviruses* **8**, 1549–1560 (1992).

15.	Rambaut, A., Lam, T. T., Max Carvalho, L. & Pybus, O. G. Exploring the temporal structure of heterochronous sequences using TempEst (formerly Path-O-Gen). *Virus Evol* **2**, vew007 (2016).

16.	Sagulenko, P., Puller, V. & Neher, R. A. TreeTime: Maximum-likelihood phylodynamic analysis. *Virus Evolution* **4**, (2018).

17.	Poppiel, R. R., Cherubin, M. R., Novais, J. J. M. & Demattê, J. A. M. Soil health in Latin America and the Caribbean. *Commun Earth Environ* **6**, 1–11 (2025).

18.	Copernicus Climate Change Service. Agrometeorological indicators from 1979 up to 2019 derived from reanalysis. ECMWF https://doi.org/10.24381/CDS.6C68C9BB (2019).

19.	Global terrestrial Human Footprint maps for 1993 and 2009 | Scientific Data. https://www.nature.com/articles/sdata201667.

20.	Earth Science Data Systems, N. Gridded Population of the World, Version 4 (GPWv4): Population Density, Revision 11 | NASA Earthdata. Earth Science Data Systems, NASA (2024).

21.	Tang, F. H. M. *et al.* CROPGRIDS: a global geo-referenced dataset of 173 crops. *Sci Data* **11**, 413 (2024).

22.	ICC | Caliper - Statistical Classifications in a Linked Open World | Food and Agriculture Organization of the United Nations. *CALIPER* https://www.fao.org/statistics/caliper/classifications/icc/en.
